# Single versus Combination Treatment in Tinnitus: An International, Multicentre, Parallel-arm, Superiority, Randomised Controlled Trial

**DOI:** 10.1101/2024.01.09.24300978

**Authors:** Stefan Schoisswohl, Laura Basso, Jorge Simoes, Milena Engelke, Berthold Langguth, Birgit Mazurek, Jose Antonio Lopez-Escamez, Dimitrios Kikidis, Rilana Cima, Alberto Bernal-Robledano, Benjamin Boecking, Jan Bulla, Christopher R. Cederroth, Holger Crump, Sam Denys, Alba Escalera-Balsera, Alvaro Gallego-Martinez, Silvano Gallus, Hazel Goedhart, Leyre Hidalgo-Lopez, Carlotta M. Jarach, Hafez Kader, Michael Koller, Alessandra Lugo, Steven C. Marcrum, Nikos Markatos, Juan Martin-Lagos, Marta Martinez-Martinez, Nicolas Muller-Locatelli, Patrick Neff, Uli Niemann, Patricia Perez-Carpena, Rüdiger Pryss, Clara Puga, Paula Robles-Bolivar, Matthias Rose, Martin Schecklmann, Tabea Schiele, Miro Schleicher, Johannes Schobel, Myra Spiliopoulou, Sabine Stark, Susanne Staudinger, Alexandra Stege, Beat Toedtli, Ilias Trochidis, Vishnu Unnikrishnan, Evgenia Vassou, Nicolas Verhaert, Carsten Vogel, Zoi Zachou, Winfried Schlee

## Abstract

Tinnitus is associated with a variety of aetiologies, phenotypes, and underlying pathophysiological mechanisms, and available treatments have limited efficacy. A combination of treatments, addressing various aspects of tinnitus, might provide a viable and superior treatment strategy.

In this international multicentre, parallel-arm, superiority, randomised controlled trial, patients with chronic subjective tinnitus were recruited from five clinical sites across the EU as part of the interdisciplinary collaborative UNITI project. Patients were randomly assigned using a web-based system, stratified by their hearing and distress level, to single or combination treatment of 12 weeks. Cognitive-behavioural therapy, hearing aids, structured counselling, and sound therapy were administered either alone or as a combination of two treatments resulting in ten treatment arms. The primary outcome was the difference in the change from baseline to week 12 in the total score of the Tinnitus Handicap Inventory (THI) between single and combination treatments in the intention-to-treat population. All statistical analysis were performed blinded to treatment allocation.

674 patients of both sexes aged between 18 and 80 years were screened for eligibility. 461 participants (190 females) with chronic subjective tinnitus and at least mild tinnitus handicap were enrolled, 230 of which were randomly assigned to single and 231 to combination treatment. Least-squares mean changes from baseline to week 12 were -11.7 for single treatment (95% confidence interval [CI], -14.4 to -9.0) and -14.9 for combination treatments (95% CI, -17.7 to -12.1), with a statistically significant group difference (*p*=0.034). Cognitive-behavioural therapy and hearing aids alone had large effect sizes, which could not be further increased by combination treatment. No serious adverse events occurred.

In this trial involving patients with chronic tinnitus, all treatment arms showed improvement in THI scores from baseline to week 12. Combination treatments showed a stronger clinical effect than single treatment, however, no clear synergistic effect was observed when combining treatments. We observed rather a compensatory effect, where a more effective treatment offsets the clinical effects of a less effective treatment.

ClinicalTrials.gov Identifier: NCT04663828.

## Main

Tinnitus is defined as “the conscious awareness of a tonal or composite noise for which there is no identifiable external acoustic source”,^1^ with an estimated prevalence of 14.4% (95% confidence interval [CI], 12.6 to 16.5) in the global population, with 2.3% (95% CI, 1.7 to 3.1) being severely affected.^2^ Severe tinnitus is associated with emotional stress, cognitive dysfunction, and/or autonomic arousal, leading to maladaptive behavioural changes and functional disability.^1^

Numerous causes and risk factors for tinnitus have been identified,^3^ whereby peripheral and central mechanisms are involved in its emergence and maintenance, exemplified by pathological alterations in the ear, along the auditory pathway^4^, as well as in non-auditory brain regions.^5^ There is a broad spectrum of aetiologies, phenotypes, and underlying pathophysiological mechanisms of tinnitus. Many adults with chronic tinnitus report having tried multiple tinnitus treatments before finding a treatment that reduces their tinnitus distress.^6^ Despite the availability of treatment guidelines,^7,8^ clear guidance on which treatment strategy is best for the individual patient is not yet available. A viable option for clinical management could be the combination of different treatment options to target various facets of this symptom simultaneously.

However, studies on the efficacy of combining clinical interventions are scarce.^9–11^ A prominent example of combining different treatment types is represented by the combination of a specific acoustic therapy with directive counselling termed Tinnitus Retraining Therapy.^12^

The primary objective of the current trial was to compare the effect of single against combination treatments for patients with chronic tinnitus. Four established treatment strategies were selected: cognitive-behavioural therapy (CBT), hearing aids (HA), structured counselling (SC), and sound therapy (ST).^13^ Participants were randomised either to a single treatment out of this set of treatments or to a combination of two treatments. Further, we attempt to overcome methodological weaknesses^14^ of previous trials by investigating a large multinational sample of tinnitus patients, using harmonised patient selection and screening procedures, as well as standardised interventions and assessments.

## Methods

### Study design

This was an investigator-initiated, international, multicentre, parallel-arm, superiority, randomised controlled clinical trial conducted in five hospitals across four European countries (Belgium, Germany, Greece, and Spain; see **Table S1** in the Supplementary Appendix) as part of the UNITI project (Unification of Treatments and Interventions for Tinnitus Patients).^15^ Included patients received treatment between April 2021 and December 2022. Detailed information about the trial rationale, design, methodological approaches, and statistical analysis strategies are published in the study protocol and statistical analysis plan (SAP).^16,17^ The study was approved by local ethics committees at every clinical site independently. Further, all authors vouch for the completeness and correctness of the data, adherence of the trial to the study protocol,^16^ as well as adherence of data analysis strategies to the SAP.^17^ A detailed list of author contributions can be found in the Supplementary Appendix. Written informed consent was obtained from all eligible patients prior to trial participation. For the preparation of this report we used the CONSORT guidelines (Consolidated Standards of Reporting Trials).^18^

### Participants

Adults of both sexes (self-reported) aged between 18 and 80 years with chronic subjective tinnitus (lasting for six months or more) were recruited and screened at each clinical site. Inclusion criteria for trial participation were at least mild tinnitus handicap according to the Tinnitus Handicap Inventory^19^ (THI; score ≥ 18) and tinnitus as primary complaint. Exclusion criteria were: presence of a mild cognitive impairment according to the Montreal Cognitive Assessment^20^ (MoCa; score ≤ 22); any relevant ear disorders or acute infections of the ear; one deaf ear; severe hearing loss (inability to communicate properly) as well as serious internal, neurological, or psychiatric conditions. Existing drug therapies with psychoactive substances had to be stable, and no start of any other tinnitus-related treatment in the last three months before trial participation was allowed. A detailed list of all eligibility criteria can be found in the trial protocol.^16^ Written informed consent was obtained from all participants.

### Randomisation and blinding

After successful on-site screening, eligible participants were stratified in four equally sized strata based on their THI total score (low [< 48] and high [≥ 48] tinnitus-related handicap) and hearing aid indication (yes and no, criteria for hearing aid indication: **Table S2)**. Participants were then randomised to one of ten treatment arms comprised of single (CBT, HA, SC, ST) and combination interventions (CBT+HA, CBT+SC, CBT+ST, HA+SC, HA+ST, SC+ST). Patients from the two strata without hearing aid indication were not randomised in treatment groups that comprised HA treatment. Randomisation was conducted at each clinical site with an interactive web response system developed together with biostatisticians from the contract research organization Excelya (www.excelya.com). Excelya was further responsible to monitor all randomisation proceedings. Treatment codes were used to assure blindness of the statistical analysis team to the type of treatment patients received. Unblinding was conducted after analyses completion. See study protocol and statistical analysis plan for more detailed information.^16,17^

### Procedures

Single and combination treatments were applied over a 12-week treatment phase. CBT was based on the concept of fear-avoidance using exposure therapy^21^ and delivered by trained psychologists or psychotherapists in weekly face-to-face group sessions (1.5-2 hours; group size: six to eight participants). For HA treatment, behind-the-ear hearing instruments (Type Signia Pure 312 7X; Sivantos Pte. Ltd., Singapore, Republic of Singapore/ WSAudiology, Lynge, Denmark) were fitted bilaterally with all noise-related signal processing deactivated by audiologists or HA acousticians according to the National Acoustic Laboratories-Non-Linear 2 generic amplification proceeding.^22^ SC and ST were self-administered on a daily basis via dedicated mobile applications.^23^ SC consisted of 12 chapters featuring structured patient education and tips on how to handle tinnitus distress. ST included a set of various artificial and naturalistic sounds. All treatment procedures were designed by dedicated experts in their respective fields (see **Table S3**) and described in detail in the study protocol.^16^ Demographic and clinical characteristics were assessed at baseline (before treatment) using the European School of Interdisciplinary Tinnitus Research Screening Questionnaire (ESIT-SQ).^24^ Outcome measures were assessed at baseline, interim (after 6 weeks of treatment), final (after 12-week treatment period), and follow-up (36 weeks after baseline) visits. An additional follow-up visit (48 weeks after baseline) was conducted on a voluntary basis. Due to its voluntary nature and the associated large amount of missing data, this additional follow-up was not included in the final outcome measure analysis.

### Outcome measures

The primary outcome was the difference in change from baseline to final visit (after 12 weeks of treatment) in the Tinnitus Handicap Inventory (THI),^19^ which consists of 25 items to quantify tinnitus handicap (total scores range: 0-100), between single and combination treatment. Changes from baseline to interim visit, and follow-up were examined in secondary analyses as well. Secondary outcome measures included the Tinnitus Functional Index (TFI),^25^ the Mini Tinnitus Questionnaire (Mini-TQ),^26^ the Patient Health Questionnaire for Depression (PHQ-D/PHQ-9),^27^ the abbreviated version of the World Health Organisation - Quality of Life questionnaire (WHO-QoL)^28^ as well as numeric rating scales (NRS) for tinnitus impairment, tinnitus loudness, tinnitus-related discomfort, annoyance, unpleasantness, and ability to ignore the tinnitus.^29^ Clinical improvement was measured with the Clinical Global Impression Scale – Improvement (CGI-I).^30^

Adverse (AE) and serious adverse events (SAE) were defined according to the guidelines for Good Clinical Practice §3 (6,8). AEs were assessed and recorded during each visit with respect to start and end date, intensity, relation to intervention, impact on treatment, and actions taken. Any SAE during the 12-week treatment phase led to a stop of the patient’s respective treatment and was immediately reported to the local ethics committee.

### Statistical Analysis

The sample size was determined a priori on an estimated effect size of 0.26, an alpha level of 5% and a power of 80% (two-sided test). Based on that, the necessary sample size is 468. Considering potential dropouts, the aim was to recruit a total sample size of N = 500.^16^

The statistical analysis was performed in the intention-to-treat (ITT) population of N = 461, including all randomised participants, regardless of compliance with the study protocol. For the primary analysis (combination against single treatments), we estimated that with a two-tailed alpha level of less than 0.05, the sample size of N = 461 provides the trial with 90% power to detect an effect size of 0.30 (lower end of 95% CI for effect size of behavioural therapy interventions according to the latest Cochrane Review on tinnitus).^31^

For the ITT analysis, missing values (THI: 18%, education: 3.5%, PHQ-9 baseline: 2.6%) were imputed using multilevel imputation (R package `mitml`)^32,33^; see **Figure S2** for the distribution of imputed THI values. The missing at random assumption was checked as described in the SAP and no indications of violation were found. As sensitivity analysis, a per-protocol analysis was conducted on all patients who met the requirements for treatment compliance as defined in the SAP (N = 185, **Table S32**).^34^ An additional sensitivity analyses was performed without imputation of the primary outcome (**Table S34**).

The analysis of the primary objective was performed in the ITT population to test the efficacy of combination treatments against single treatments. Further comparisons between single versus combination treatments for all 4 single treatments separately (CBT single vs. combined, HA single vs. combined, SC single vs. combined, ST single vs. combined) as well as comparisons between all ten treatment arms were performed.^16,34^ To address all objectives, mixed effect models were applied (with REML using the `lme4` R package)^35^ by considering the outcome as the response variable and including the corresponding objective, time point (baseline, interim visit, final visit, and follow-up), and objective-by-time interaction as fixed effects, including centre and subject ID as random intercepts. The models were adjusted for the following covariates: age, sex, educational attainment, hearing aid indication, and PHQ-9 baseline scores.^34^ The results of the remaining objectives as described in the SAP are reported in the Supplementary Appendix.

Results are reported as least-squares mean changes (obtained via the `emmeans` R package)^36^ with 95% confidence intervals. All analyses were performed in R (version 4.2.2).

De-identified data (pseudo-anonymised code) were gathered in a central database, which was regularly monitored and systematically checked for missing and invalid data (every six weeks). After database closure and prior to analysis, data from each clinical centre were checked again for validity and completeness. This study was registered at ClinicalTrials.gov, NCT04663828.

## Results

Between Apr 16, 2021, and Sept 20, 2022, 674 persons with tinnitus were assessed for eligibility, of whom 461 (68.3%) fulfilled the inclusion criteria and consented to participate. After randomisation, 230 were allocated to single treatments and 231 were allocated to combination treatments (**Figure 1**). The initial planned sample size for the trial was 500 patients.^16^ Since our study plan required a recruitment of an exact number of patients with specific tinnitus profiles (eligibility criteria and stratification proceedings), plus the trial was performed during the COVID-19 pandemic, recruitment and inclusion processes took us significantly longer than expected. Hence, we closed the trial in December 2022 with N = 461 included and treated patients, in order to keep to the schedule of our funding period. A post hoc power computation indicates that with a two-tailed alpha level of less than 5%, the available sample size of N = 461 provides our trial with 79.5% power to detect an effect size of 0.26.

**Figure 1.**
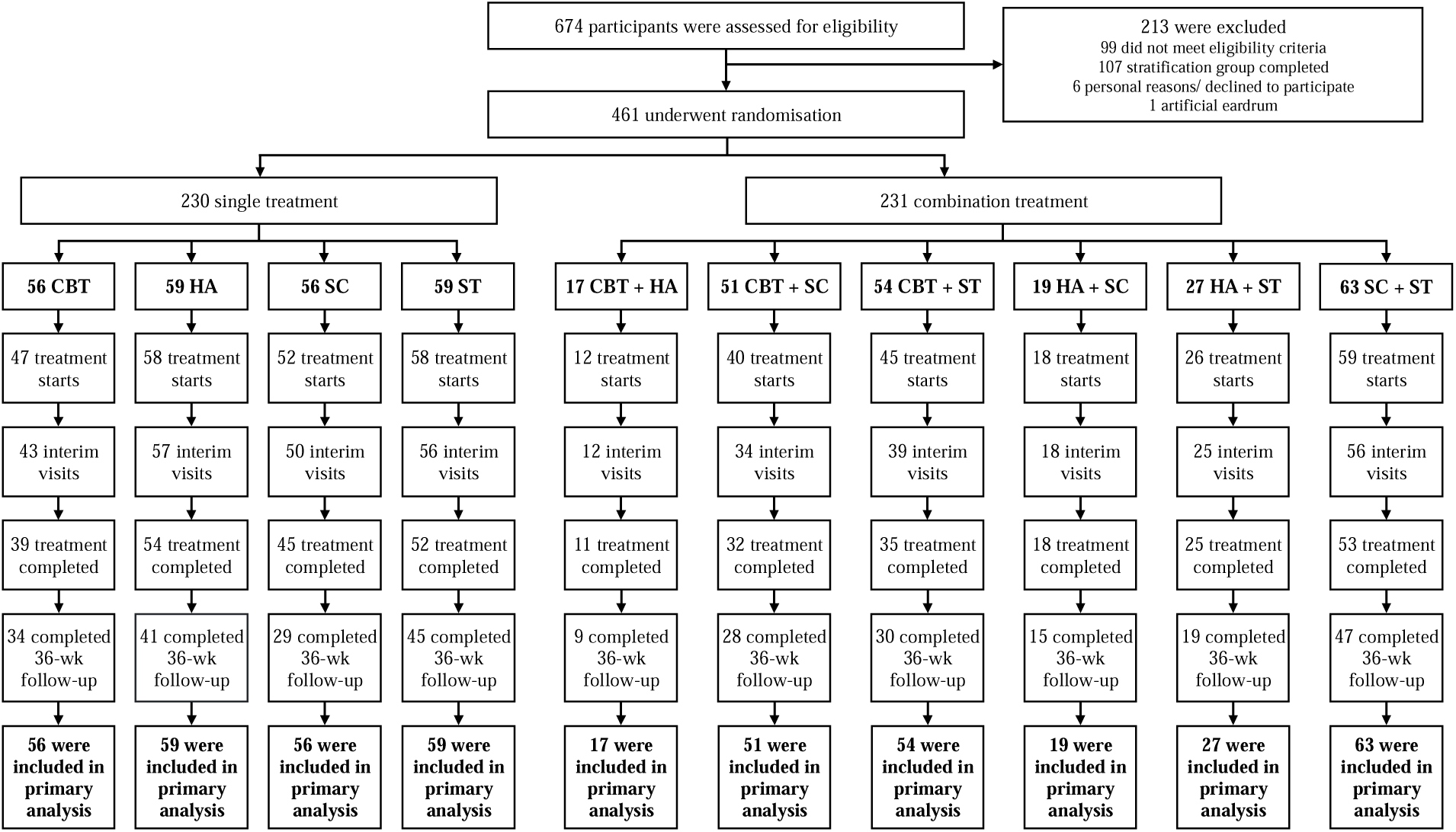
Trial Profile. A total of 674 patients were screened, of whom 461 met the trial inclusion criteria and were randomly assigned to one of ten treatment arms comprised of a single treatment or a combination of two treatments out of four different therapy approaches - cognitive-behavioural therapy (CBT), hearing aids (HA), structured counselling (SC), and sound therapy (ST). 230 (49.9%) were assigned to single treatments (CBT, HA, SC, or ST) and 231 (50.1%) were assigned to combination treatments (CBT+HA, CBT+SC, CBT+ST, HA+SC, HA+ST, SC+ST). Patients without hearing aid indication were only randomised to treatments without HA. An extended version of the patient’s flowchart can be found in **Figure S1**. Quantity and reasons for trial exclusion during eligibility assessments and trial discontinuation can be seen from **Tables S5 – S9**.

**Table 1** shows the baseline characteristics by treatment arm. Mean baseline THI total scores were 48.5 (SD 19.5) in the single treatment group and 47.4 (SD 19.9) in the combined treatment group. Except for age and hearing aid indication, the baseline characteristics were generally well balanced between the treatment arms (see **Table 1** and **Table S10**). Both age and hearing aid indication were considered as covariates during statistical analyses. The difference in hearing aid indication results from randomising only individuals with relevant hearing loss to HA treatment arms. Results of audiometric measurements are shown in **Figure S3** and **S4**. Participants’ baseline characteristics were similar to the group of persons with tinnitus seeking medical help in the general population (**Table S4**).

**Table 1.**
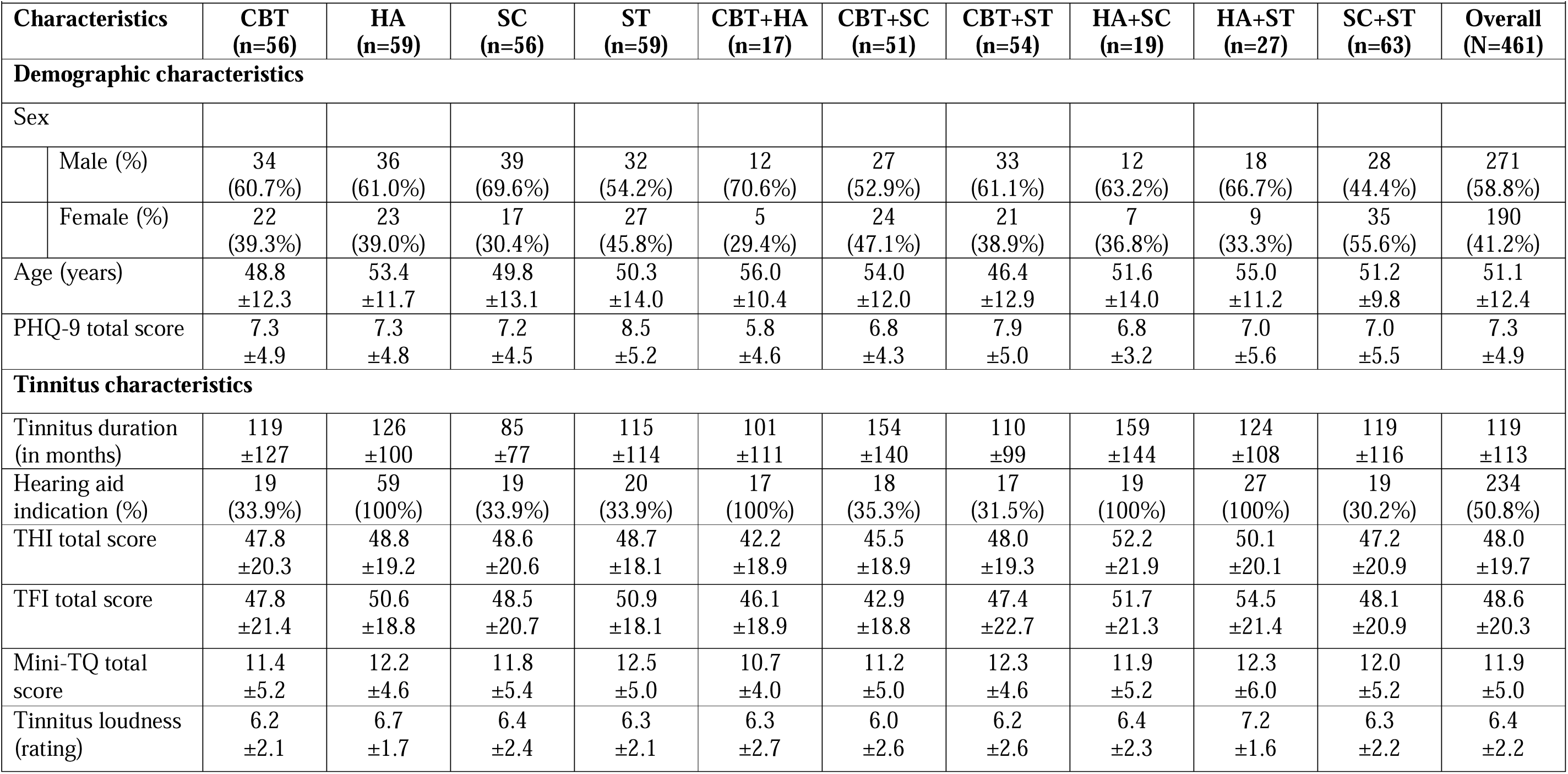
Demographic and Clinical Characteristics of the Participants at Baseline. Data are n (%) or mean ± SD. PHQ-9 scores range from 0 to 27, with higher scores indicating greater severity of depression. The definition for hearing aid indication is given in Table S2. THI scores range from 0 to 100, with higher scores indicating greater severity of tinnitus. TFI scores range from 0 to 100, with higher scores indicating greater severity of tinnitus. Mini-TQ scores range from 0 to 24, with higher scores indicating greater severity of tinnitus. Tinnitus loudness (rating) scores range from 0 to 10, with higher scores indicating greater loudness of tinnitus. Abbreviations: CBT = Cognitive-Behavioural Therapy; HA = Hearing Aids; PHQ-9 = Patient Health Questionnaire for Depression; SC = Structured Counselling; ST = Sound Therapy; TFI = Tinnitus Functional Index; THI = Tinnitus Handicap Inventory; TQ = Tinnitus Questionnaire.

Regarding the primary objective, the least-squares mean change from baseline to week 12 in the THI total score was -11.7 (95% confidence interval [CI], -14.4 to -9.0) for the single treatment groups and -14.9 (95% CI, -17.7 to -12.1) for the combination treatment arms (see **Figure 2 & Table 2)** (interaction effect [single vs. combination treatments at final visit vs. baseline] *ß* = 3.2, 95% CI, 0.2 to 6.1, *p* = 0.034). Model parameters and model assumptions for the primary objective can be found in **Table S12** and **Figure S5**. The least-squares mean change from baseline to week 12 in the THI total score for the single vs. combination treatment comparison for each treatment strategy is reported in **Table 2**, and separately for every treatment arm in **Table 3** and **Figure S6**; and further separated by hearing aid indication in **Table S13** and tinnitus severity in **Table S14**. **Figure 2** shows least-squares mean changes from baseline to interim visit at week 6, final visit at week 12, and follow-up at week 36 for both the overall and individual single-combination treatment comparison. The results of the remaining objectives (as outlined in the SAP)^17^ and time points (interim visit and follow up) are reported in **Tables S16 – S18**.

**Figure 2.**
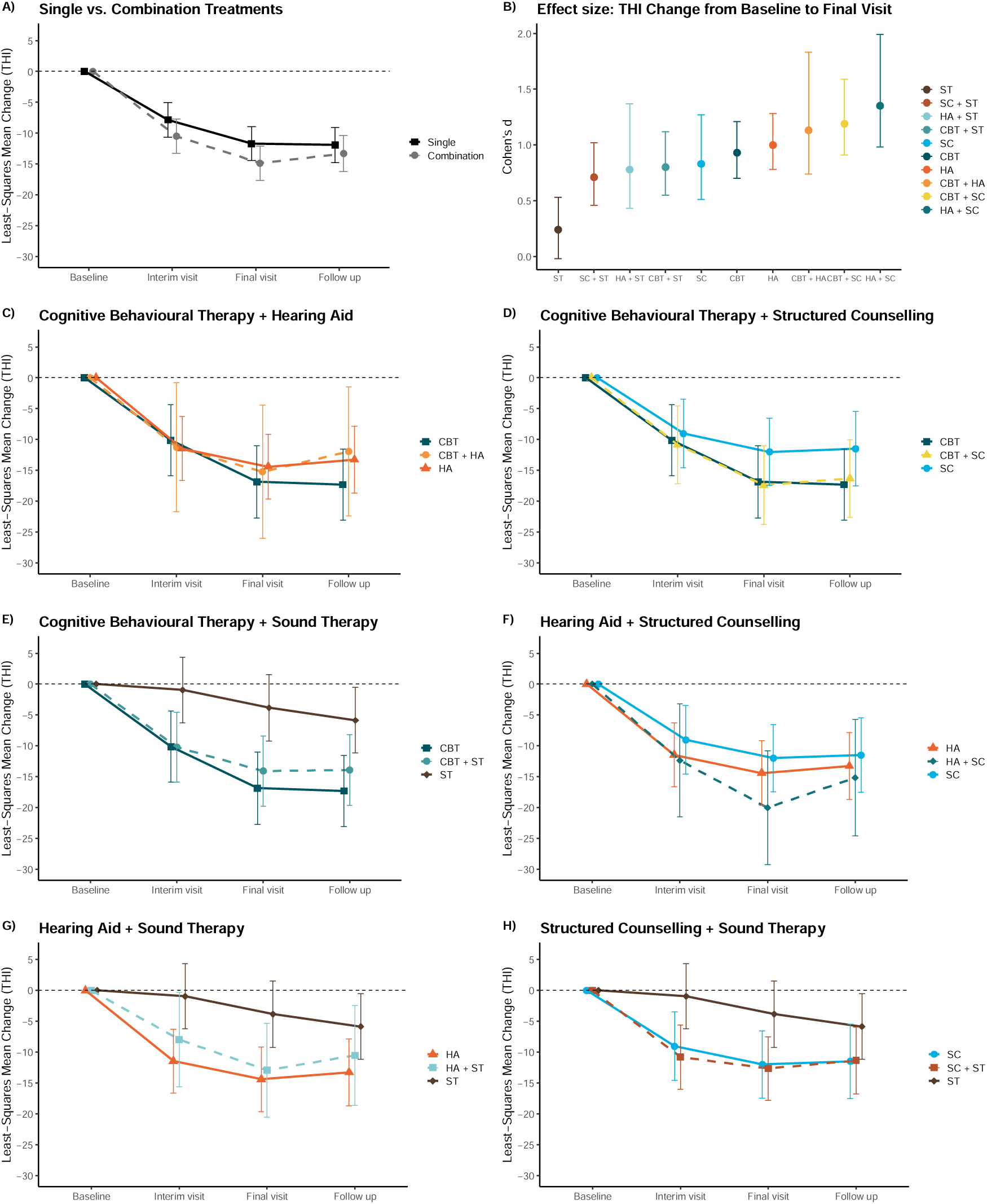
Least-Squares Mean Changes from Baseline to interim visit (6w), final visit (12w) and follow-up (36w) in THI total score. **A)** single and combination treatments; **C)** CBT+HA; **D)** CBT+SC; **E)** CBT+ST; **F)** HA+SC; **G)** HA+ST; **H)** SC+ST; and **B)** Cohen’s d values for all treatment arms (change in THI total score from baseline to final visit). Total THI scores range from 0 to 100, with higher scores indicating greater severity of tinnitus. Error bars represent 95% confidence intervals. Abbreviations: CBT = Cognitive-Behavioural Therapy; HA = Hearing Aids; SC = Structured Counselling; ST = Sound Therapy; THI = Tinnitus Handicap Inventory.

**Table 2.**
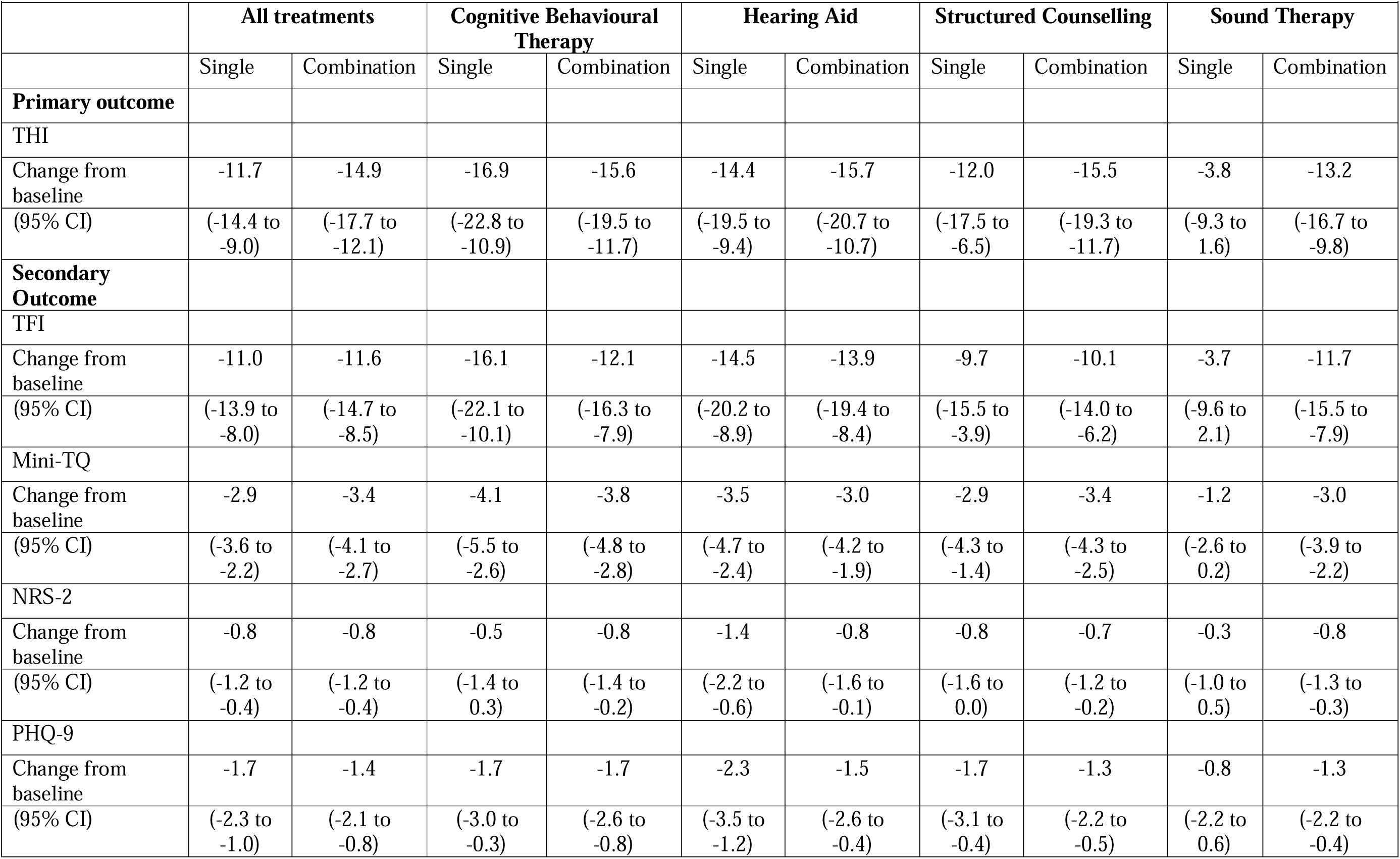
Primary and Secondary Clinical Outcomes at Final Visit: Single vs. Combination (ITT). Values depict least-squares mean changes at week 12 for primary and secondary outcomes with 95% confidence intervals. Higher total scores on the THI, TFI and Mini-TQ indicate greater severity of tinnitus. Higher total scores on the NRS-2 indicate greater loudness of tinnitus. Higher total scores on the PHQ-9 indicate greater severity of depression. Further objectives and secondary clinical outcomes not reported in this table can be seen in the Supplementary Appendix. Abbreviations: NRS = Numeric Rating Scale; PHQ-9 = Patient Health Questionnaire for Depression; TFI = Tinnitus Functional Index; THI = Tinnitus Handicap Inventory; TQ = Tinnitus Questionnaire.

| **Table 2. Primary and Secondary Clinical Outcomes at Final Visit: Single vs. Combination (Intention-to-Treat Population).** |  |  |  |  |  |  |  |  |  |  |
| --- | --- | --- | --- | --- | --- | --- | --- | --- | --- | --- |
|  | **All treatments** |  | **Cognitive Behavioural Therapy** |  | **Hearing Aid** |  | **Structured Counselling** |  | **Sound Therapy** |  |
|  | Single | Combination | Single | Combination | Single | Combination | Single | Combination | Single | Combination |
| **Primary outcome** |  |  |  |  |  |  |  |  |  |  |
| THI |  |  |  |  |  |  |  |  |  |  |
| Change from baseline | -11.7 | -14.9 | -16.9 | -15.6 | -14.4 | -15.7 | -12.0 | -15.5 | -3.8 | -13.2 |
| (95% CI) | (-14.4 to -9.0) | (-17.7 to -12.1) | (-22.8 to -10.9) | (-19.5 to -11.7) | (-19.5 to -9.4) | (-20.7 to -10.7) | (-17.5 to -6.5) | (-19.3 to -11.7) | (-9.3 to 1.6) | (-16.7 to -9.8) |
| **Secondary Outcome** |  |  |  |  |  |  |  |  |  |  |
| TFI |  |  |  |  |  |  |  |  |  |  |
| Change from baseline | -11.0 | -11.6 | -16.1 | -12.1 | -14.5 | -13.9 | -9.7 | -10.1 | -3.7 | -11.7 |
| (95% CI) | (-13.9 to -8.0) | (-14.7 to -8.5) | (-22.1 to -10.1) | (-16.3 to -7.9) | (-20.2 to -8.9) | (-19.4 to -8.4) | (-15.5 to -3.9) | (-14.0 to -6.2) | (-9.6 to 2.1) | (-15.5 to -7.9) |
| Mini-TQ |  |  |  |  |  |  |  |  |  |  |
| Change from baseline | -2.9 | -3.4 | -4.1 | -3.8 | -3.5 | -3.0 | -2.9 | -3.4 | -1.2 | -3.0 |
| (95% CI) | (-3.6 to -2.2) | (-4.1 to -2.7) | (-5.5 to -2.6) | (-4.8 to -2.8) | (-4.7 to -2.4) | (-4.2 to -1.9) | (-4.3 to -1.4) | (-4.3 to -2.5) | (-2.6 to 0.2) | (-3.9 to -2.2) |
| NRS-2 |  |  |  |  |  |  |  |  |  |  |
| Change from baseline | -0.8 | -0.8 | -0.5 | -0.8 | -1.4 | -0.8 | -0.8 | -0.7 | -0.3 | -0.8 |
| (95% CI) | (-1.2 to -0.4) | (-1.2 to -0.4) | (-1.4 to 0.3) | (-1.4 to -0.2) | (-2.2 to -0.6) | (-1.6 to -0.1) | (-1.6 to 0.0) | (-1.2 to -0.2) | (-1.0 to 0.5) | (-1.3 to -0.3) |
| PHQ-9 |  |  |  |  |  |  |  |  |  |  |
| Change from baseline | -1.7 | -1.4 | -1.7 | -1.7 | -2.3 | -1.5 | -1.7 | -1.3 | -0.8 | -1.3 |
| (95% CI) | (-2.3 to -1.0) | (-2.1 to -0.8) | (-3.0 to -0.3) | (-2.6 to -0.8) | (-3.5 to -1.2) | (-2.6 to -0.4) | (-3.1 to -0.4) | (-2.2 to -0.5) | (-2.2 to 0.6) | (-2.2 to -0.4) |

**Table 3.** Primary and Secondary Clinical Outcomes at Final Visit: All treatment Arms (ITT). Values depict least-squares mean changes at week 12 for primary and secondary outcomes with 95% confidence intervals. Higher total scores on the THI, TFI and Mini-TQ indicate greater severity of tinnitus. Higher total scores on the NRS-2 indicate greater loudness of tinnitus. Higher total scores on the PHQ-9 indicate greater severity of depression. Cohens d indicate the standardised effect size of the respective treatment. The effect sizes and the corresponding confidence intervals were first computed in each of the 50 imputed data sets before they were averaged to a single value. Further objectives and secondary clinical outcomes not reported in this table can be seen in the Supplementary Appendix. Abbreviations: CBT = Cognitive-Behavioural Therapy; HA = Hearing Aids; NRS = Numeric Rating Scale; PHQ-9 = Patient Health Questionnaire for Depression; SC = Structured Counselling; ST = Sound Therapy; TFI = Tinnitus Functional Index; THI = Tinnitus Handicap Inventory; TQ = Tinnitus Questionnaire.

|  | CBT | HA | SC | ST | CBT+HA | CBT+SC | CBT+ST | HA+SC | HA+ST | SC+ST |
| --- | --- | --- | --- | --- | --- | --- | --- | --- | --- | --- |
| **Primary outcome** |  |  |  |  |  |  |  |  |  |  |
| THI |  |  |  |  |  |  |  |  |  |  |
| Change from baseline | -16.9 | -14.4 | -12.0 | -3.8 | -15.2 | -17.4 | -14.1 | -20.0 | -12.9 | -12.7 |
| (95% CI) | (-22.7 to -11.0) | (-19.7 to -9.2) | (-17.5 to -6.5) | (-9.2 to 1.5) | (-26.0 to -4.4) | (-23.8 to -11.0) | (-19.8 to -8.4) | (-29.3 to -10.8) | (-20.5 to -5.3) | (-17.8 to -7.5) |
| Cohen's d<br>(95% CI) | 0.93<br>(0.70 to 1.21) | 1.00<br>(0.78 to 1.28) | 0.83<br>(0.51 to 1.27) | 0.24<br>(-0.02 to 0.53) | 1.13<br>(0.74 to 1.83) | 1.19<br>(0.91 to 1.59) | 0.80<br>(0.55 to 1.12) | 1.35<br>(0.98 to 1.99) | 0.78<br>(0.43 to 1.37) | 0.71<br>(0.46 to 1.02) |
| **Secondary Outcome** |  |  |  |  |  |  |  |  |  |  |
| TFI |  |  |  |  |  |  |  |  |  |  |
| Change from baseline | -16.1 | -14.5 | -9.7 | -3.7 | -15.1 | -10.9 | -12.2 | -10.1 | -15.8 | -9.4 |
| (95% CI) | (-22.2 to -10.0) | (-20.1 to -8.9) | (-15.6 to -3.8) | (-9.5 to 2.0) | (-26.1 to -4.0) | (-17.4 to -4.4) | (-18.5 to -5.9) | (-20.0 to -0.2) | (-24.0 to -7.6) | (-15.1 to -3.8) |
| Mini-TQ |  |  |  |  |  |  |  |  |  |  |
| Change from baseline | -4.1 | -3.5 | -2.9 | -1.2 | -4.0 | -4.1 | -3.6 | -3.2 | -2.3 | -2.9 |
| (95% CI) | (-5.5 to -2.6) | (-4.8 to -2.2) | (-4.3 to -1.4) | (-2.6 to 0.2) | (-6.7 to -1.3) | (-5.6 to -2.6) | (-5.0 to -2.2) | (-5.5 to -0.9) | (-4.3 to -0.4) | (-4.2 to -1.6) |
| NRS-2 |  |  |  |  |  |  |  |  |  |  |
| Change from baseline | -0.5 | -1.4 | -0.8 | -0.3 | -1.0 | -0.9 | -0.7 | -0.3 | -1.1 | -0.7 |
| (95% CI) | (-1.4 to 0.3) | (-2.1 to -0.6) | (-1.6 to 0.0) | (-1.1 to 0.5) | (-2.5 to 0.6) | (-1.8 to 0.0) | (-1.5 to 0.1) | (-1.6 to 1.1) | (-2.2 to -0.1) | (-1.5 to 0.0) |
| PHQ-9 |  |  |  |  |  |  |  |  |  |  |
| Change from baseline | -1.7 | -2.3 | -1.7 | -0.9 | -1.2 | -1.8 | -1.8 | -2.0 | -1.3 | -0.8 |
| (95% CI) | (-3.0 to -0.3) | (-3.6 to -1.1) | (-3.1 to -0.4) | (-2.2 to 0.4) | (-3.7 to 1.2) | (-3.2 to -0.3) | (-3.2 to -0.4) | (-4.2 to 0.2) | (-3.1 to 0.6) | (-2.1 to 0.4) |

Regarding the secondary outcome measures, least-squares mean change from baseline to week 12 for TFI, Mini-TQ, PHQ-9, WHO-QoL, and NRS (all objectives) are shown in **Tables 2** and **3** as well as **Tables S19 – S31**. Results of CGI-I are reported descriptively for single and combination treatment groups at final visit, see **Figure S7 & S8**, and separated by hearing aid indication (**Figure S9**) and tinnitus severity (**Figure S10**).

No serious adverse event was evident in any participant. Adverse events appeared in 49 (21.3%) participants in single treatment groups, and in 49 (21.2%) participants in combination treatment groups. A full listing of all adverse events is provided in **Table S11**. Information on treatment adherence is given in **Figure S1**.

Pairwise post-hoc contrasts for the THI least-squares mean change revealed statistically significant (Bonferroni adjusted) differences between ST and CBT, ST and CBT+SC, ST and CBT+ST, ST and HA, and ST and HA+SC. For all other treatment contrasts, no statistically significant differences were found (all p-values > 0.050). Statistical parameters for all post- hoc contrasts are listed in **Table S15.** The intention-to-treat and the sensitivity analysis yielded similar results (**Table S34**). Per-protocol findings were different for the overall single vs. combination contrast (no statistical superiority of combination treatment; *ß* = 2.8, 95% CI, -1.6 to 7.2, *p* = 0.206) (**Figure S11, Tables S32 – S33**). Exploratory analysis included the effect size estimates Cohen’s d for all treatment arms which are shown in **Table 3** and **Figure 2**.

## Discussion

In this randomised trial on chronic tinnitus, the efficacy of established tinnitus treatments (CBT, HA, SC, and ST) applied either alone or as a combination of two treatments was investigated. All treatments were safe and the improvement in THI scores from baseline to week 12 was statistically stronger for combination compared to single treatment. However, a more detailed analysis of our data by pairwise post hoc comparisons of the various treatment arms suggests that the additional effect of a treatment combination depends on the efficacy of a single treatment. In the case of ST, a clear superiority in favour of combination treatment was present, with the combination CBT+ST being statistically more effective than single ST. Importantly, there was no statistically significant difference between CBT alone and CBT+ST. This finding shows that combining a treatment with low efficacy (in this case ST) together with a treatment of high efficacy (in this case CBT) does not lead to a simple regression to the mean.

Rather the high-efficacy treatment counterbalances the effect of the low-efficacy treatment and elevates the clinical improvement up to a level comparable to the single high-efficacy treatment. Together with the observation that ST was the treatment which demonstrated the smallest improvements in tinnitus-related handicap (statistically significant less than CBT, HA, CBT+SC, CBT+ST, HA+SC), the additional beneficial effect of a treatment combination appears to depend on how effective a single treatment already performs. For the single treatment arm with ST, we observed a weak effect size of 0.24 (CI, -0.02 to 0.53) while combinations of treatments including ST yielded medium to strong effect sizes: SC+ST (Cohen’s d = 0.71, CI, 0.46 to 1.02), HA+ST (Cohen’s d = 0.78, CI, 0.43 to 1.37), and CBT+ST (Cohen’s d = 0.80. CI, 0.55 to 1.12), which is driven by the combination treatments of higher efficacy.

The weak clinical efficacy of sound treatment alone is in line with previous work where sound treatment was used as an active comparator.^37^ This trial shows that combining a treatment of weak clinical efficacy with a treatment of stronger clinical efficacy counterbalanced the weak effect and provokes a clinical improvement comparable to the stronger effect. On the other hand, if a single treatment is already effective, a combination might not result in a synergistic effect.

Previous investigations evaluated combination treatments for tinnitus as well.^9–11^ For instance it was demonstrated that Tinnitus Retraining Therapy,^12^ which combines a specific acoustic therapy with directive counselling, reduced tinnitus symptoms more effectively than counselling alone.^9^

However, this is the first systematic trial to investigate CBT, HA, ST, and SC within the scope of one investigation. With the present trial, we can directly put into perspective the effect size of CBT as the most established treatment in tinnitus,^7,8,31^ with HA, ST, and SC (ST and SC provided with mobile applications) as well as their combinations as treatment options for tinnitus. The combination of HA+SC, which provided the strongest effect size in our trial, has not been investigated so far, and data about the clinical efficacy in tinnitus are not yet available.^38,39^

For the interpretation of the results, it should be considered that we worked with a selected set of four tinnitus treatments and combinations of two treatment types. Thus, it remains unknown, whether the combination of other treatment sets or combinations of three or more treatment types would lead to additional treatment benefits. The duration of treatment was 12 weeks in all treatment arms. Meaningful clinical improvements were observed in most treatment arms after 6 weeks and improved further towards the final assessment after 12 weeks and remained during the follow-up period.

Despite the usage of interventions allowing for a high level of patient flexibility (SC and ST via mobile applications, HA), treatment compliance/adherence was low (see **Table S35**) and drop-out rates were high in our trial (PP sample of 185 patients). With the application of two treatments in combination, the chances that one or even both treatments are not conducted as intended are increasing. Further, high drop-out rates are a well-known issue in mobile health interventions.^40^ Another reason could be that patients were randomized to treatments and did not receive the treatment they desired. Under ideal treatment compliance/adherence (PP analysis), we observed no overall superiority of combination treatments.

A potential explanation for this incongruency between ITT and PP analysis might be that under perfect conditions (PP), a single treatment which is conducted properly is already effective on its own and thus there is no clear additional beneficial effect of a combination treatment. However, if one or two treatments are not properly conducted (ITT), as it is most probably the case in the everyday clinical treatment of tinnitus, a combination of treatments provides an additional benefit. Our results indicate that there is a high need for further research to better understand the clinical effects of combination treatment; to get more profound insights behind the reasons for low treatment adherence; and in approaches to increase treatment adherence in daily clinical practice, such as the implementation of behavioral change techniques or more extensive patient education.

A placebo group was not included in this trial, as the answer to the main question (comparison of single and combined treatment) did not require a placebo arm. Nevertheless, a placebo group may have been helpful as an anchor for comparison with the ten treatment arms. However, our results of CBT as single treatment correspond very well to meta-analytic data of its efficacy^31^ and thus provide an anchor for a well-established evidence based treatment approach. Further, our data demonstrates low efficacy of ST as a single treatment, supporting its use as an active control condition in randomised controlled trials.^41^ Thus, the two treatment arms CBT and ST can be considered as reliable reference anchors for the interpretation of the results of the other 8 investigated treatment arms.

Even though in 18% of all participants data of the primary outcome was missing, the sensitivity analysis (no imputation of THI) came to a similar finding regarding our primary objective.

In this trial involving adults with chronic tinnitus, we found that 12 weeks of treatment with CBT, HAs, SC, or ST applied as single or in combinations of two treatments led to an amelioration in tinnitus-related handicap. There was no unambiguous synergistic effect of treatment combination, rather a compensatory effect, where a more effective treatment offsets the clinical effects of a less effective treatment. In clinical situations where it is unclear which treatment will benefit the patient, a combination of treatments might help to increase the chances of treatment success.

## Data availability

De-identified data generated from this trial will initially be available only on request for researchers to reproduce results, later publicly via ZENODO. Current data availability will be reported on the UNITI website (https://uniti.tinnitusresearch.net/).

## Contributions

Stefan Schoisswohl (SSch) contributed to conceptualisation, investigation, formal analysis, methodology, project administration, supervision, visualisation, and writing – original draft; Winfried Schlee (WS) contributed to conceptualisation, funding acquisition, formal analysis, methodology, project administration, supervision, visualisation, and writing – original draft; Berthold Langguth (BL) contributed to conceptualisation, funding acquisition, methodology, supervision, and writing – review & editing; Laura Basso (LB) and Milena Engelke (ME) contributed to data curation, formal analysis, methodology, visualisation, and writing – original draft; Rüdiger Pryss (RP) contributed to funding acquisition, investigation, methodology, software, and writing – review & editing; Birgit Mazurek (BM), Jose Antonio Lopez-Escamez (JALE), Dimitrios Kikidis (DK), and Rilana Cima (RC) contributed to conceptualisation, funding acquisition, methodology, and writing – review & editing; Myra Spiliopoulou (MSp) contributed to funding acquisition, data curation, formal analysis, and writing – review & editing; Susanne Staudinger (SStau) contributed to conceptualisation, project administration, investigation, and writing – review & editing; Carsten Vogel (CV) contributed to investigation, methodology, software, and writing – review & editing; Jorge Simoes (JS), Uli Niemann (UN), Clara Puga (CP), Miro Schleicher (MSchl), Carlotta M. Jarach (CMJ), Hafez Kader (HK), and Vishnu Unnikrishnan (VU) contributed to data curation, formal analysis, methodology, and writing – review & editing; Benjamin Boecking (BB) and Martin Schecklmann (MSche) contributed to conceptualisation, methodology, investigation, and writing – review & editing; Christopher R. Cederroth (CRC) contributed to funding acquisition, resources, and writing – review & editing; Steven C. Marcrum (SCM) contributed to conceptualisation, investigation, and writing – review & editing; Patrick Neff (PN) contributed to conceptualisation, methodology, and writing – review & editing; Johannes Schobel (JS) contributed to investigation, software, and writing – review & editing; Alberto Bernal-Robledano (ABR), Marta Martinez-Martinez (MMM), Nicolas Muller- Locatelli (NML), Patricia Perez-Carpena (PPC), Paula Robles-Bolivar (PRB), Matthias Rose (MR), Tabea Schiele (TS), Sam Denys (SD), Alba Escalera-Balsera (AEB), Alvaro Gallego-Martinez (AGM), Leyre Hidalgo-Lopez (LHL), Nikos Markatos (NM), Juan Martin-Lagos (JML), Sabine Stark (SSta), Alexandra Stege (AS), Evgenia Vassou (EV), Nicolas Verhaert (NV), and Zoi Zachou (ZZ) contributed to investigation and writing – review & editing; Jan Bulla (JB) and Beat Toedtli (BT) contributed to formal analysis, validation, and writing – review & editing; Silvano Gallus (SG) contributed to funding acquisition and writing – review & editing; Michael Koller (MK), Hazel Goedhart (HG) and Holger Crump (HC) contributed to conceptualisation and writing – review & editing; and Alessandra Lugo (AL) and Ilias Trochidis (IT) contributed to writing – review & editing.

All authors had full access to all the data in the study and had final responsibility for the decision to submit for publication. SSch, WS, LB, and ME have directly accessed and verified the underlying data reported in the manuscript.

## Funding

This clinical trial received funding from the European Union’s Horizon 2020 Research and Innovation Program (grant agreement number: 848261). The funders had no influence on trial design and had no role in the collection, analysis, interpretation of the data, preparation of the manuscript or in the decision to submit the manuscript for publication.

## Competing interests

SSch received funding outside the present study from dtec.bw – Digitalization and Technology Research Center of the Bundeswehr (MEXT project). dtec.bw is funded by the European Union – NextGenerationEU. ME received research funding outside the present study from the Rainwater Charitable Foundation and the Sonova Holding AG. BL received consulting fees outside the present study from Schwabe Pharma AG, Neuromod, Sea Pharma, and Rovi; payment or honoraria for lectures, presentations, speakers bureaus, manuscript writing or educational events outside the present study from Schwabe and Neuromod; payment for expert testimony outside the present study from the Bavarian State; has an unpaid leadership or fiduciary role outside the present study in the German Society for Brain Stimulation in Psychiatry and the Tinnitus Research Initiative; owns stock or stock options form Sea Pharma; and received equipment outside the present study from Neurocare and Daymed. BM received support for attending meetings and/ or travel outside the present study from the Charité – Universitätsmedizin Berlin. JALE received support for attending meetings and conferences outside the present study from the University of Sydney. RC received support for attending meetings and conferences outside the present study from the KU Leuven University, Faculty of Psychology and Educational Sciences, Health Psychology, Belgium and Adelante Center of Expertise in Rehabilitation and Audiology, The Netherlands. AEB received research funding outside the present study from ibs.Granada/Fundación para la Investigación Biosanitaria de Andalucía Oriental (FIBAO), European Molecular Biology Organisation (EMBO) and Centro de Investigación Biomédica en Red Enfermedades Raras (CIBERER). AGM received research funding outside the present study from the CECEU 2020, Andalusian Government of Spain (grant number: DOC_01677), and support for attending meetings and/ or travel from the Andalusian Health Department (Grant number: PI- 0266-2021 – GEN4PHEN). PPC received research funding outside the present study from the Consejería de Salud y Familias, Junta de Andalucía. 2020, Contrato Posdoctorales Especialistas (RH–0150–2020), and the Instituto de Salud Carlos III., Bases neurofisiológicas y perfil de seguridad de la terapia sonora en pacientes con acúfeno crónico severo (PI22/01838). RP is a stakeholder of the Lenox uG. The Lenox uG also holds shares of the HealthStudyClub GmbH. MSp received payment or honoraria for lectures, presentations, speaker bureaus, manuscript writing or educational events outside the present study from VAIA (Vlaamse AI Academie: https://www.vaia.be/en/), which is a collaboration between all the universities and universities for applied sciences and arts in Flanders; payment effected by KU Leuven. NV received research funding outside the present study from the Flanders Research Foundation and VLAIO - KU Leuven, and has a planned patent with KU Leuven. WS is a stakeholder of the Lenox uG. Lenox uG also holds shares of the HealthStudyClub GmbH; received consulting fees outside the present study from Pansatori GmbH and Pohl-Boskamp GmbH & Co. KG; received payment or honoraria for lectures, presentations, speaker bureaus, manuscript writing or educational events outside the present study from Schwabe Pharma AG and Medical Tribune; has a patent planned, issued or pending (US20120046713A1).

## Ethical approval

The study was approved by local ethics committees of Granada, Athens, Leuven, Regensburg and Berlin (combined ethical approval for German sites).

## Acknowledgments

We would like to thank all patients who participated in this trial, without whom this research would not have been possible. We would like to further thank the whole consortium of the UNITI-project for their feedback and support. Moreover, we would like to thank Simon Grund for his support regarding the `mitml` R package.

## Notes

### Competing Interest Statement

Details on competing interest can be found in the preprint.

### Clinical Trial

NCT04663828

### Clinical Protocols

https://trialsjournal.biomedcentral.com/articles/10.1186/s13063-021-05835-z

https://trialsjournal.biomedcentral.com/articles/10.1186/s13063-023-07303-2

### Funding Statement

This clinical trial received funding from the European Unions Horizon 2020 Research and Innovation Program (grant agreement number 848261)

### Author Declarations

The study was independently approved by the following ethics committees: University of Regensburg, Regensburg, Germany (combined ethical approval for clinical sites Berlin and Regensburg); Katholieke Universiteit Leuven, Leuven, Belgium; Ethniko Kai Kapodistriako Panepistimo Athinon, Athens, Greece; Hospital Universitario Virgen de las Nieves/ Hospital Clinico Universitario San Cecilio, Granada, Spain.

### Summary of Updates

Discussion was extended. Abstract was updated.

