## Supplementary Appendix for "Single versus Combination Treatment in Tinnitus: An International, Multicentre, Parallel-arm, Superiority, Randomised Controlled Trial" (from another version of this preprint: version 2, 2024-02-26)

**Table Of Contents for Supplementary Appendix**

1. ***Table S1: Clinical Sites and Principal Investigators***

| **Country** | **Clinical Site** | **Principal Investigator** |
| --- | --- | --- |
| Belgium | Katholieke Universiteit Leuven, Leuven | Asst. Prof. Dr. Rilana Cima  Prof. Dr. Johan Vlaeyen |
| Germany | Tinnitus Center, Charité – Universitatsmedizin Berlin, corporate member of Freie Universität Berlin, Humboldt-Universität zu Berlin, and Berlin Institute of Health, Berlin | Prof. Dr. Birgit Mazurek  Dr. Benjamin Boecking |
|  | University of Regensburg, Regensburg  (RCT coordinator) | Prof. Dr. Berthold Langguth  Prof. Dr. Martin Schecklmann  Dr. Stefan Schoisswohl |
| Greece | Ethniko Kai Kapodistriako Panepistimo Athinon, Athens, | PD Dr. Dimitris Kikidis  Assoc. Prof. Dr. Athanasios Bibas |
| Spain | Hospital Universitario Virgen de las Nieves, Granada  Hospital Clinico Universitario San Cecilio, Granada | PD. Dr. Jose Antonio López-Escámez  Dr. Juan Manuel Espinosa-Sanchez  Dr. Patricia Perez Carpena |

2. ***Author Contributions***

| **Author name** | **Contribution** | **Author name** | **Contribution** |
| --- | --- | --- | --- |
| Stefan Schoisswohl, Ph.D. | Conceptualisation; investigation; formal analysis; methodology; project administration; supervision; visualisation; writing – original draft | Marta Martinez-Martinez, M.D., Ph.D. | Investigation; writing – review & editing |
| Laura Basso, Ph.D. | Data curation; formal analysis; methodology; visualisation; writing – original draft | Nicolas Muller-Locatelli, M.D. | Investigation; writing – review & editing |
| Jorge Simoes, Ph.D. | Data curation; formal analysis; methodology; writing – review & editing | Patrick Neff, Ph.D. | Conceptualisation; Methodology; writing – review & editing |
| Milena Engelke, M.Sc. | Data curation; formal analysis; methodology; visualisation; writing – original draft | Uli Niemann, Dr. Ing. | Data curation; formal analysis; methodology; writing – review & editing |
| Berthold Langguth, M.D. | Conceptualisation; funding acquisition; methodology; supervision; writing – review & editing | Patricia Perez-Carpena, M.D., Ph.D. | Investigation; writing – review & editing |
| Birgit Mazurek, M.D., Ph.D | Conceptualisation; funding acquisition; methodology; writing – review & editing | Rüdiger Pryss, Ph.D. | Funding acquisition; investigation; methodology; software; writing – review & editing |
| Jose Antonio Lopez-Escamez, M.D., Ph.D. | Conceptualisation; funding acquisition; methodology; writing – review & editing | Clara Puga, M.Sc. | Data curation; formal analysis; methodology; writing – review & editing |
| Dimitrios Kikidis, M.D., Ph.D. | Conceptualisation; funding acquisition; methodology; writing – review & editing | Paula Robles-Bolivar, M.Sc. | Investigation; writing – review & editing |
| Rilana Cima, Ph.D. | Conceptualisation; funding acquisition; methodology; writing – review & editing | Matthias Rose, Ph.D. | Investigation; writing – review & editing |
| Alberto Bernal-Robledano, M.Sc. | Investigation; writing – review & editing | Martin Schecklmann, Ph.D. | Conceptualisation; investigation; methodology; writing – review & editing |
| Benjamin Boecking, Ph.D., DClinPsy. | Conceptualisation; methodology; investigation; writing – review & editing | Tabea Schiele, M.Sc. | Investigation; writing – review & editing |
| Jan Bulla, Ph.D. | Formal analysis; writing – review & editing | Miro Schleicher, M.Sc. | Data curation; formal analysis; methodology; writing – review & editing |
| Christopher R. Cederroth, Ph.D. | Funding acquisition; resources; writing – review & editing | Johannes Schobel, Ph.D. | Investigation; software; writing – review & editing |
| Sam Denys, Ph.D. | Investigation; writing – review & editing | Myra Spiliopoulou, Dr. habil. | Funding acquisition; data curation, formal analysis; writing – review & editing |
| Alba Escalera-Balsera, M.Sc. | Investigation; writing – review & editing | Sabine Stark, Dipl.-Psych. | Investigation; writing – review & editing |
| Alvaro Gallego-Martinez, Ph.D. | Investigation; writing – review & editing | Susanne Staudinger, M.A. | Conceptualisation; project administration; investigation; writing – review & editing |
| Silvano Gallus, Ph.D. | Funding acquisition; writing – review & editing | Alexandra Stege, Ph.D. | Investigation; writing – review & editing |
| Leyre Hidalgo-Lopez, M.D. | Investigation; writing – review & editing | Beat Toedtli, Ph.D. | Formal analysis; validation; writing – review & editing |
| Carlotta M. Jarach, M.Sc. | Data curation; formal analysis; methodology; writing – review & editing | Ilias Trochidis, M.Sc. | writing – review & editing |
| Hafez Kader, M.Sc. | Data curation; formal analysis; methodology; writing – review & editing | Vishnu Unnikrishnan, M.Sc. | Data curation; formal analysis; methodology; writing – review & editing |
| Michael Koller, Ph.D. | Conceptualisation; writing – review & editing | Evgenia Vassou, M.Sc. | Investigation; writing – review & editing |
| Alessandra Lugo, Ph.D. | Writing – review & editing | Nicolas Verhaert, M.D, Ph.D. | Investigation; writing – review & editing |
| Steven C. Marcrum, Ph.D. | Conceptualisation; investigation; writing – review & editing | Carsten Vogel, M.Sc. | Investigation; methodology; software; writing – review & editing |
| Nikos Markatos, B.Sc. (Honors) | Investigation; writing – review & editing | Zoi Zachou, M.D. | Investigation; writing – review & editing |
| Juan Martin-Lagos, M.D. | Investigation; writing – review & editing | Winfried Schlee. Ph.D. | Conceptualisation; funding acquisition; formal analysis; methodology; project administration; supervision; visualisation; writing – original draft |

*Note.* All authors fulfilled the necessary requirement for authorship and agreed to be fully accountable for all aspects of the present work. All authors read and approved the final manuscript.

3. ***Table S2: Criteria for HA indication***

| **Frequency (Hz)** | **Minimum threshold (dB HL)** | **Maximum threshold (dB HL)** |
| --- | --- | --- |
| 125 | 10 | 60 |
| 250 | 10 | 60 |
| 500 | 20 | 65 |
| 1000 | 25 | 70 |
| 2000 | 35 | 75 |
| 3000 | 40 | 80 |
| 4000 | 45 | 80 |
| 6000 | 45 | 80 |

*Note.* Minimum and maximum threshold (dB HL) per frequency used for the definition of HA indication. HA=Hearing aid.

4. ***Table S3: Responsible Persons for the Conception of Treatments***

|  | **Profession** |
| --- | --- |
| **Cognitive Behavioural Therapy** | |
| Boecking, B. | Psychologist, Psychotherapist |
| Cima, R. | Psychologist, Psychotherapist |
| Schecklmann, M. | Psychologist, Psychotherapist |
| **Hearing Aid** | |
| Dettling-Papargyris, J. | Hearing aid acoustician |
| Kikidis, D. | Audiologist |
| Marcrum, S. C. | Audiologist |
| Mazurek, B. | ENT-Physician |
| Oppel, K. | Hearing aid acoustician |
| Schiele, T. | Doctoral student |
| Schlee, W. | Psychologist |
| **Structured Counselling** | |
| Engelke, M. | Psychologist |
| Schlee, W. | Psychologist |
| **Sound Therapy** | |
| Neff, P. | Psychologist |
| Schlee, W. | Psychologist |

5. ***Table S4: Representativeness of Study Participants***

| **Category** | **Example** |
| --- | --- |
| Condition under investigation | Chronic tinnitus. The Tinnitus persisted for a minimum of 6 months. |
| Sex | No sex effect was found for the primary outcome (THI) when comparing single versus combination treatments for chronic tinnitus. 41·2 % of the participants were female, 58·8% were male. |
| Age | No age effect was found for the primary outcome (THI) when comparing single versus combination treatments for chronic tinnitus.  The average age was 51·1 years. In a study by Probst et al. 2017, it was reported that the majority of patients visiting the specialised tinnitus clinic in Regensburg are in the age range of 45 - 55 years. |
| Race or ethnic group | Race or ethnicity was not assessed. No conclusions can be drawn about the influence of race or ethnicity. |
| Geography | The 5 centres were located in four European countries: Germany (2), Greece, Netherlands, Spain. Conclusions about countries with dissimilar health care systems are limited. |
| Severity | The average THI score at baseline was 48·0, which is considered moderate tinnitus distress. Participants with slight or no distress at screening (THI < 18), were excluded from the study. |
| Other considerations | In this trial, we compared single versus combination treatment of four clinical intervention types typically used for tinnitus treatment: cognitive behavioural therapy, hearing aids, sound therapy, and structured counselling for a duration of 12 weeks. The treatments were implemented in a typical way. Conclusions about treatments with different implementations are limited. |
| Overall representativeness of this trial | This trial showed that combining a treatment of weak clinical efficacy with a treatment of stronger clinical efficacy leads to an improvement of the overall clinical outcome.  We recognise that the trial was limited to only four different clinical intervention types and their combinations. We do not know whether other treatment combinations, which are not part of the trial, would be superior to their respective singe treatments.  We cannot conclude about combination treatments with three or more clinical interventions.  All treatments were implemented in a typical way; however, it would be possible to implement them differently or apply the treatment for a shorter or longer time. We do not know whether other implementation would have the same results. |

6. ***Table S5: Excluded Patients from Study Participation***

|  | **N** |
| --- | --- |
| **Not meeting inclusion criteria** |  |
| Primary complaint tinnitus | 2 |
| Chronic tinnitus (≥ six months) | 1 |
| Age between 18 and 80 years | 2 |
| A score of ≥ 18 in the Tinnitus Handicap Inventory (THI; Newman et al., 1996) – at least mild tinnitus distress | 32 |
| A score of > 22 in the Montreal Cognitive Assessment (MoCa; Nasreddine et al., 2005) – absence of mild cognitive impairment | 14 |
| Ability and willingness to use the UNITI mobile applications on smartphones | 4 |
| Openness to use a HA (if indication and allocation to HA group) | ·· |
| Ability to understand and consent to the research (hearing ability, intellectual capacity) | ·· |
| Ability to participate in all relevant visits (no plans for e.g., long-term holidays or pregnancy) | ·· |
| Existing drug therapies with psychoactive substances (e.g., antidepressants or anticonvulsants) must be stable for at least 30 days at the beginning of the therapeutic intervention. The drug therapy should remain constant during the course of the study. Necessary changes do not constitute an exclusion criterion per se, but need to be recorded. | ·· |
| **Meeting exclusion criteria** |  |
| Objective tinnitus or heartbeat-synchronous tinnitus as primary complaint | 7 |
| Otosclerosis / acoustic neuroma or other relevant ear disorders with fluctuation hearing | 10 |
| Present acute infections (acute otitis media, otitis externa, acute sinusitis) | 1 |
| Meniere's disease or similar syndromes (but not vestibular migraine) | 2 |
| Serious internal, neurological or psychiatric conditions | 4 |
| Epilepsy or other disorders of the central nervous system (e.g., brain tumor or encephalitis) | 1 |
| Clinically relevant drug, medication or alcohol abuse up to 12 weeks before study start | ·· |
| Severe hearing loss – inability to communicate properly in the course of the study | 11 |
| One deaf ear | 4 |
| Missing written informed consent | 1 |
| Start of any other tinnitus related treatments, especially hearing aids, structured counselling,  sound therapy (with special devices; expecting long term effects) or cognitive behavioural therapy in the last 3 months before the start of the study | 3 |
| **Other reasons** |  |
| Stratification group completed | 107 |
| Personal reasons/ declined to participate | 6 |
| Artificial eardrum | 1 |
| **Total number of excluded patients** | 213 |

7. ***Table S6: Quantity and Reasons of Drop-outs before Treatment Start per Treatment Arm***

| **Treatment arm** | **N** | **Reasons (N)** |
| --- | --- | --- |
| CBT | 9 | No time (2); unable to travel to clinic (2); rejected CBT (2); no internet connection (1); started another treatment (1); interested in HA (1) |
| HA | 1 | Declined to participate (1) |
| SC | 4 | Rejected SC (1); interested in HA (1); declined to participate (1); app treatment not started (1) |
| ST | 1 | No capacity for daily app usage (1) |
| CBT + HA | 5 | Unable to travel to clinic (1); rejected HA and CBT (1); No capacity for CBT (1); suspected Morbus Meniere (1); lost (1) |
| CBT + SC | 11 | No time (4); lost (2); financial reasons (1); job reasons (1); chronic fatigue syndrome (1); rejected CBT (1); rejected CBT & app treatment not started (1) |
| CBT + ST | 9 | Rejected CBT + ST (3); no time (3); lost (1); personal reasons (1); job reasons (1) |
| HA + SC | 1 | Unexpected personal problems (1) |
| HA + ST | 1 | Rejected HA (1) |
| SC + ST | 4 | Interested in HA (1); no time (1); suspected acoustic neuroma (1); app treatment not started (1) |
| Total number of drop-outs before treatment start | 46 |  |

*Note.* CBT = Cognitive-Behavioural Therapy; HA = Hearing Aids; SC = Structured Counselling; ST = Sound Therapy.

***Table S7: Quantity and Reasons of Drop-outs during Treatment per Treatment Arm – before Interim Visit***

| **Treatment arm** | **N** | **Reasons (N)** |
| --- | --- | --- |
| CBT | 4 | Issues with daily app measures (1); no time (1); health worsening (1); personal reasons (1) |
| HA | 1 | Rejected HA (1) |
| SC | 2 | Not satisfied with treatment (1); lack of benefit & issues with daily app measures (1) |
| ST | 3 | Tinnitus worsening (1); tinnitus worsening & not satisfied with app treatment; no time due to illness (1) |
| CBT + HA | ·· | ·· |
| CBT + SC | 6 | Unsatisfied with CBT (1); health reasons (1); lost (1); no benefit of CBT (1); job reasons (1); rejected CBT (1) |
| CBT + ST | 6 | Tinnitus worsening (1); lost (1); no time (1); unable to travel to clinic (1); rejected CBT (1); treatment termination after first CBT session (1) |
| HA + SC | ·· | ·· |
| HA + ST | ·· | ·· |
| SC + ST | 3 | Tinnitus worsening (1); no reasons (1); lost (1) |
| Total number of drop-outs during treatment – before interim visit | 25 |  |

*Note.* CBT = Cognitive-Behavioural Therapy; HA = Hearing Aids; SC = Structured Counselling; ST = Sound Therapy.

8. ***Table S8: Quantity and Reasons of Drop-outs during Treatment per Treatment Arm – after Interim Visit***

| **Treatment arm** | **N** | **Reasons (N)** |
| --- | --- | --- |
| CBT | 4 | No time (2); unable to travel to clinic (1); job reasons (1) |
| HA | 3 | Lost (2); started another HA treatment (1) |
| SC | 5 | Tinnitus worsening (1); lost (1); lack of benefit (1); health worsening/ reasons (1); did not want to continue (1) |
| ST | 3 | Not satisfied with treatment (2); lost (1) |
| CBT + HA | 1 | No time (1) |
| CBT + SC | 2 | No time (1); no longer attends CBT sessions (1) |
| CBT + ST | 4 | No benefit of CBT (2); tinnitus worsening (1); lost (1) |
| HA + SC | ·· | ·· |
| HA + ST | 1 | App treatment was never started (1) |
| SC + ST | 3 | App treatment was never started (1); lost (1); did not want to come to clinic due to COVID-19 pandemic (1) |
| Total number of drop-outs during treatment – after interim visit | 26 |  |

*Note.* CBT = Cognitive-Behavioural Therapy; HA = Hearing Aids; SC = Structured Counselling; ST = Sound Therapy.

9. ***Table S9: Quantity and Reasons of Patients lost to Follow-up***

| **Treatment arm** | **N** | **Reasons (N)** |
| --- | --- | --- |
| CBT | 5 | No response (3); health reasons (1); no reasons (1) |
| HA | 5 | Job reasons (2); no response (2); no reasons (1) |
| SC | 11 | No response (6); not interested in follow-up (3); no reasons (1); health reasons (1) |
| ST | 2 | No response (1); not available for follow-up (1) |
| CBT + HA | 1 | No response (1) |
| CBT + SC | 3 | No response (1); not interested in follow-up (1); terminated follow-up too late (1) |
| CBT + ST | 1 | Deceased (1) |
| HA + SC | 1 | No response (1) |
| HA + ST | 3 | No response (3) |
| SC + ST | 4 | No response (2); wanted to discontinue (1); not available for follow-up (1) |
| Total number of patients lost to follow-up | 36 |  |

*Note.* CBT = Cognitive-Behavioural Therapy; HA = Hearing Aids; SC = Structured Counselling; ST = Sound Therapy.

10. ***Figure S1: Extended Patients Flowchart (Intention-to-treat and Per-protocol Sample)***

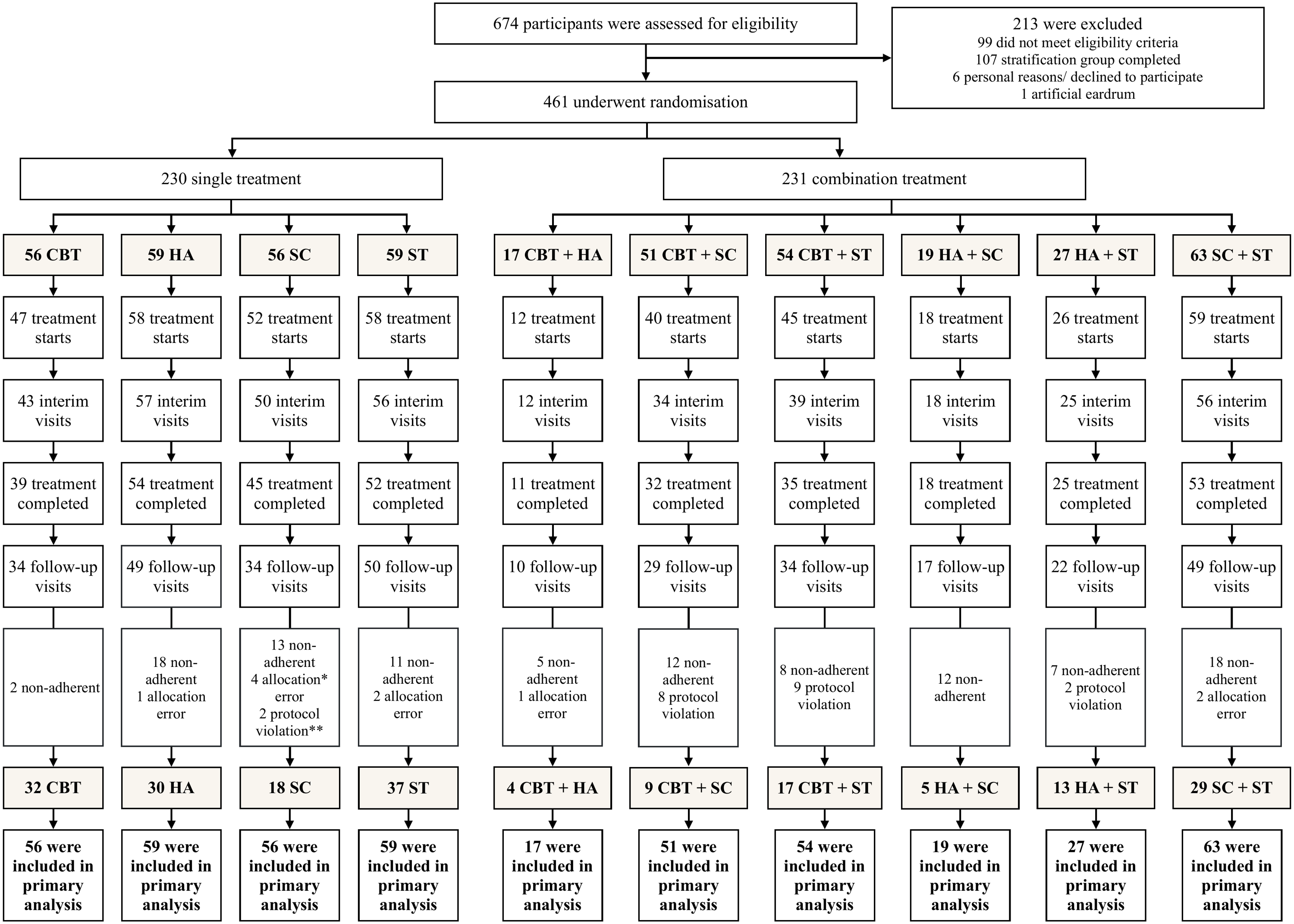

*Note.* A total of 674 patient were screened of whom 461 (ITT sample) met the trial inclusion criteria and were randomly assigned to one of ten treatment arms comprised of a single treatment or a combination of two treatments using a set of four different therapy approaches - cognitive-behavioural therapy (CBT), hearing aids (HA), structured counselling (SC), and sound therapy (ST). 230 were assigned to single treatments and 231 were assigned to combination treatments. 231 subjects were excluded during the screening process. Available data for the primary outcome (incl. missing data) per study visit and treatment arm can be seen. The number of treatment non-adherence, protocol violations and allocation errors are equally shown per treatment arm. From the sample of 194 patients for the per-protocol analysis, nine had missings in covariates relevant for the analysis, resulting in a sample of 185 patients used for the per-protocol analysis.

11. ***Table S10: Demographic and clinical characteristics of the participants at baseline (stratified by treatment arm).***

|  | **CBT (n=56)** | **HA (n=59)** | **SC (n=56)** | **ST (n=59)** | **CBT + HA (n=17)** | **CBT + SC (n=51)** | **CBT + ST (n=54)** | **HA + SC (N=19)** | **HA + ST (n=27)** | **SC + ST (n=63)** | **Total (N=461)** |
| --- | --- | --- | --- | --- | --- | --- | --- | --- | --- | --- | --- |
| **Education — no. (%)** |  |  |  |  |  |  |  |  |  |  |  |
| Elementary or middle school | 11 (21·2%) | 18 (30·5%) | 12 (21·8%) | 12 (21·4%) | 7 (43·8%) | 20 (41·7%) | 10 (20·0%) | 4 (21·1%) | 6 (22·2%) | 17 (27·0%) | 117 (26·3%) |
| High school | 10 (19·2%) | 10 (16·9%) | 12 (21·8%) | 16 (28·6%) | 2 (12·5%) | 7 (14·6%) | 11 (22·0%) | 6 (31·6%) | 6 (22·2%) | 13 (20·6%) | 93 (20·9%) |
| University | 31 (59·6%) | 31 (52·5%) | 31 (56·4%) | 28 (50·0%) | 7 (43·8%) | 21 (43·8%) | 29 (58·0%) | 9 (47·4%) | 15 (55·6%) | 33 (52·4%) | 235 (52·8%) |
| **Tinnitus Presentation (ESIT-SQ B2) — no. (%)** |  |  |  |  |  |  |  |  |  |  |  |
| constant | 48 (94·1%) | 57 (96·6%) | 49 (92·5%) | 51 (94·4%) | 13 (86·7%) | 44 (93·6%) | 45 (93·8%) | 18 (100%) | 26 (100%) | 57 (90·5%) | 408 (94·0%) |
| intermittent | 3 (5·88%) | 2 (3·39%) | 4 (7·55%) | 3 (5·56%) | 2 (13·3%) | 3 (6·38%) | 3 (6·25%) | 0 (0%) | 0 (0%) | 6 (9·52%) | 26 (5·99%) |
| **Tinnitus Loudness [dB] (Tinnitus Matching)** |  |  |  |  |  |  |  |  |  |  |  |
| Mean (SD) | 35·3 (21·6) | 42·0 (21·1) | 32·8 (23·0) | 37·4 (23·1) | 48·7 (18·6) | 37·9 (19·1) | 37·4 (18·6) | 44·9 (27·6) | 51·4 (19·9) | 38·0 (21·8) | 38·8 (21·7) |
| Median [Min, Max] | 37·3 [1, 83] | 46·0 [0, 82·5] | 30·0 [2·5, 88] | 35·0 [5, 90] | 48·0 [20, 78] | 40·5 [0, 75] | 35·0 [5, 85] | 41·3 [0, 100] | 53·8 [10, 95] | 35·0 [2·50, 92] | 40·0 [0, 100] |
| **Residual inhibition:  Both ears — no. (%)** |  |  |  |  |  |  |  |  |  |  |  |
| none | 17 (70·8%) | 10 (41·7%) | 11 (45·8%) | 16 (50·0%) | 3 (42·9%) | 14 (56·0%) | 17 (53·1%) | 5 (50·0%) | 8 (57·1%) | 17 (48·6%) | 118 (52·0%) |
| partially | 5 (20·8%) | 10 (41·7%) | 8 (33·3%) | 8 (25·0%) | 2 (28·6%) | 5 (20·0%) | 7 (21·9%) | 4 (40·0%) | 5 (35·7%) | 7 (20·0%) | 61 (26·9%) |
| complete | 2 (8·33%) | 4 (16·7%) | 5 (20·8%) | 8 (25·0%) | 2 (28·6%) | 6 (24·0%) | 8 (25·0%) | 1 (10·0%) | 1 (7·14%) | 11 (31·4%) | 48 (21·1%) |
| **Residual inhibition: Left ear — no. (%)** |  |  |  |  |  |  |  |  |  |  |  |
| none | 9 (50·0%) | 10 (62·5%) | 9 (56·3%) | 10 (71·4%) | 2 (66·7%) | 7 (63·6%) | 7 (70·0%) | 3 (75·0%) | 1 (16·7%) | 4 (33·3%) | 62 (56·4%) |
| partially | 5 (27·8%) | 2 (12·5%) | 1 (6·25%) | 2 (14·3%) | 0 (0%) | 2 (18·2%) | 1 (10·0%) | 1 (25·0%) | 3 (50·0%) | 6 (50·0%) | 23 (20·9%) |
| complete | 4 (22·2%) | 4 (25·0%) | 6 (37·5%) | 2 (14·3%) | 1 (33·3%) | 2 (18·2%) | 2 (20·0%) | 0 (0%) | 2 (33·3%) | 2 (16·7%) | 25 (22·7%) |
| **Residual inhibition: Right ear — no. (%)** |  |  |  |  |  |  |  |  |  |  |  |
| none | 9 (60·0%) | 12 (63·2%) | 11 (78·6%) | 10 (90·9%) | 5 (83·3%) | 10 (66·7%) | 6 (75·0%) | 3 (60·0%) | 3 (60·0%) | 6 (54·5%) | 75 (68·8%) |
| partially | 2 (13·3%) | 4 (21·1%) | 1 (7·14%) | 0 (0%) | 0 (0%) | 1 (6·67%) | 1 (12·5%) | 2 (40·0%) | 0 (0%) | 3 (27·3%) | 14 (12·8%) |
| complete | 4 (26·7%) | 3 (15·8%) | 2 (14·3%) | 1 (9·09%) | 1 (16·7%) | 4 (26·7%) | 1 (12·5%) | 0 (0%) | 2 (40·0%) | 2 (18·2%) | 20 (18·3%) |
| **WHO-QoL 1 score: Physical health** |  |  |  |  |  |  |  |  |  |  |  |
| Mean (SD) | 12·4 (1·73) | 12·5 (1·78) | 12·4 (1·84) | 11·8 (2·10) | 13·0 (1·62) | 12·9 (2·04) | 12·6 (1·57) | 13·1 (1·26) | 12·7 (2·11) | 12·7 (1·96) | 12·5 (1·87) |
| Median [Min, Max] | 13·0 [7, 15] | 13·0 [7, 15] | 13·0 [7, 17] | 11·5 [7, 16] | 13·0 [10, 16] | 13·0 [9, 17] | 13·0 [10, 15] | 13·0 [11, 15] | 13·0 [10, 17] | 13·0 [8, 17] | 13·0 [7, 17] |
| **WHO-QoL 2 score: Psychological health** |  |  |  |  |  |  |  |  |  |  |  |
| Mean (SD) | 13·3 (1·92) | 13·6 (1·61) | 13·5 (1·99) | 13·2 (2·02) | 13·9 (1·03) | 14·1 (1·84) | 13·7 (1·72) | 14·2 (1·48) | 13·3 (2·09) | 13·5 (1·95) | 13·6 (1·85) |
| Median [Min, Max] | 13·0 [8, 17] | 14·0 [9, 16] | 14·0 [9, 18] | 13·0 [6, 17] | 14·0 [12, 16] | 14·0 [10, 17] | 14·0 [11, 17] | 14·0 [11, 17] | 13·0 [8, 17] | 14·0 [9, 17] | 14·0 [6, 18] |
| **WHO-QoL 3 score: Social factors** |  |  |  |  |  |  |  |  |  |  |  |
| Mean (SD) | 14·5 (3·27) | 14·3 (3·15) | 14·5 (3·03) | 14·0 (3·11) | 14·6 (2·83) | 14·8 (3·01) | 14·6 (3·15) | 15·1 (2·78) | 14·6 (3·14) | 14·9 (2·96) | 14·5 (3·06) |
| Median [Min, Max] | 15·0 [8, 20] | 15·0 [5, 20] | 15·0 [8, 20] | 15·0 [4, 20] | 15·0 [9, 20] | 15·0 [8, 20] | 15·0 [9, 20] | 15·5 [9, 20] | 15·0 [8, 20] | 16·0 [7, 20] | 15·0 [4, 20] |
| **WHO-QoL 4 score: Environment** |  |  |  |  |  |  |  |  |  |  |  |
| Mean (SD) | 15·8 (2·09) | 15·9 (2·15) | 15·7 (2·59) | 15·5 (2·35) | 16·1 (1·82) | 16·4 (1·93) | 15·6 (1·92) | 15·8 (1·77) | 15·7 (2·38) | 16·0 (2·30) | 15·8 (2·18) |
| Median [Min, Max] | 16·0 [11, 20] | 16·0 [8, 20] | 16·0 [9, 20] | 15·5 [9, 20] | 16·0 [13, 19] | 16·0 [13, 20] | 16·0 [12, 20] | 15·5 [13, 19] | 16·0 [11, 20] | 16·0 [12, 20] | 16·0 [8, 20] |

*Note*. Data are n (%), mean (SD), or median [Min, Max]. Tinnitus loudness level in dB was obtained by tinnitus matching, with higher values indicating greater perceived tinnitus loudness. Residual inhibition is the brief tinnitus suppression following acoustic stimulation. WHO-QoL scores range from 4 to 20, with higher scores indicating higher quality of life in the respective domain. CBT = Cognitive-Behavioural Therapy; HA = Hearing Aids; SC = Structured Counselling; ST = Sound Therapy.

12. ***Table S11: Quantity and Type of Adverse Events per Treatment Arm***

| ***Treatment arm*** | ***Adverse event*** | ***Treatment arm*** | ***Adverse event*** |
| --- | --- | --- | --- |
| ***CBT***  ***(n = 13)*** | Reconstruction nasal septum | ***CBT + HA***  ***(n = 5)*** | Herpes zoster (thigh) |
|  | Thoracic strain |  | Skin Excision |
|  | Additional burden due to husband's temporary absence |  | Mental breakdown |
|  | Cold |  | Invasion Russia |
|  | Diverticulitis disease |  | Ear pain due to hearing aids |
|  | Torn tendon on the foot | ***CBT + SC***  ***(n = 12)*** | Vaccine reaction (Moderna) |
|  | Sickness of spouse |  | Lithotripsy |
|  | Toothache |  | Total endoprosthesis hip |
|  | Cold (suspected Covid-19 positive) |  | Leukoplakia vocal cord (surgery) |
|  | Gastrointestinal infection |  | Chronic Fatigue Syndrome |
|  | Worsening general health status |  | Vaccination reaction |
|  | Death of close friend |  | Death of the cat, depressive symptoms of the significant other |
|  | Worries about pet |  | Severe health problems of partner |
| ***HA***  ***(n = 10)*** | Mole removal |  | Angina |
|  | Acute Urine Retention |  | Death of pet |
|  | Tinnitus worsening |  | Heavy cold |
|  | Retinal tear |  | Inheritance dispute with siblings |
|  | Disease of the mother-in-law | ***CBT + ST***  ***(n = 13)*** | Common cold |
|  | Loss of mother-in-law |  | Covid-19 infection |
|  | Urinary tract infection |  | Acute pain testis (left) |
|  | Sleep problems |  | Covid-19 infection |
|  | Covid-19 infection |  | Bereavement in the family |
|  | Covid-19 infection |  | Cold |
| ***SC***  ***(n = 11)*** | Thyroid nodule (benign) |  | Mild temporary hearing loss |
|  | Aggravation of tinnitus |  | Dizziness |
|  | Diverticulitis |  | Worsening psychological health due to irritable colon or depression |
|  | Tooth extraction |  | Sleep rhythm disturbed |
|  | Septoplasty |  | Invasion Russia |
|  | Stye |  | Covid-19 infection |
|  | Professional reorientation |  | Tinnitus worsening |
|  | Parents-in-law's dementia exacerbation | ***HA + SC***  ***(n = 3)*** | Broken foot |
|  | Bicycle accident |  | Ear infection due to hearing aid |
|  | Additional task at work |  | Toothache |
|  | Shoulder pain | ***HA + ST***  ***(n = 5)*** | Inflammation of the ear |
| ***ST***  ***(n = 15)*** | Surgery hand (plate removal) |  | Frozen Shoulder |
|  | Burglary |  | Travel problem on Airport |
|  | Bicycle accident |  | Severe cold |
|  | Influenza |  | Work stress |
|  | Vaccine reaction (Covid-19) | ***SC + ST***  ***(n = 11)*** | Lymph node removal (groin) |
|  | Depression |  | Forthcoming divorce |
|  | Tooth infection and extraction |  | A lot of stress at the beginning of the school year |
|  | Great workload and stress |  | Covid-19 infection |
|  | Sprained foot |  | Migraine |
|  | Disastrous vacation |  | Tinnitus worsening |
|  | Cold |  | Tinnitus worsening |
|  | Cold |  | Accident |
|  | Persistent tiredness |  | Stress at work |
|  | Gastrointestinal infection |  | Bicycle accident |
|  | Tinnitus unchanged but more present and therefore more bothersome |  | Conflict with spouse |

*Note*. CBT = Cognitive-Behavioural Therapy; HA = Hearing Aids; SC = Structured Counselling; ST = Sound Therapy.

13. ***Figure S2: Box-and-whisker plot of observed and imputed THI values***

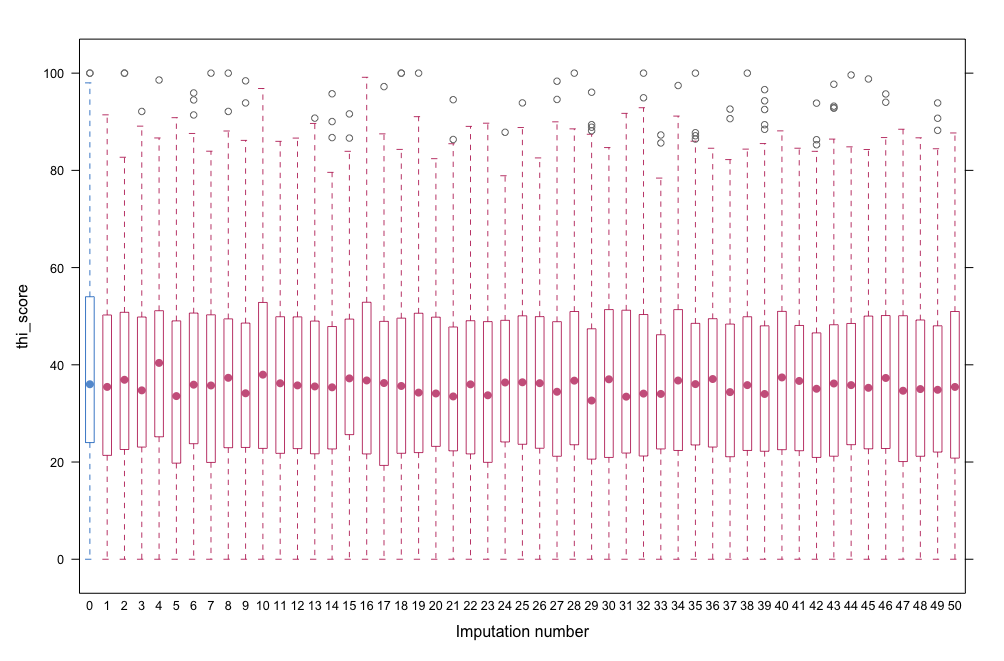

*Note*: Blue (0): Distribution of observed THI values (all visits). Red (1-50): Distribution of imputed THI values in all imputed data sets (all visits). THI = Tinnitus Handicap Inventory.

14. ***Figure S3: Audiogram in Patients With and Without Hearing Aid Indication for Single and Combined Treatment Groups***

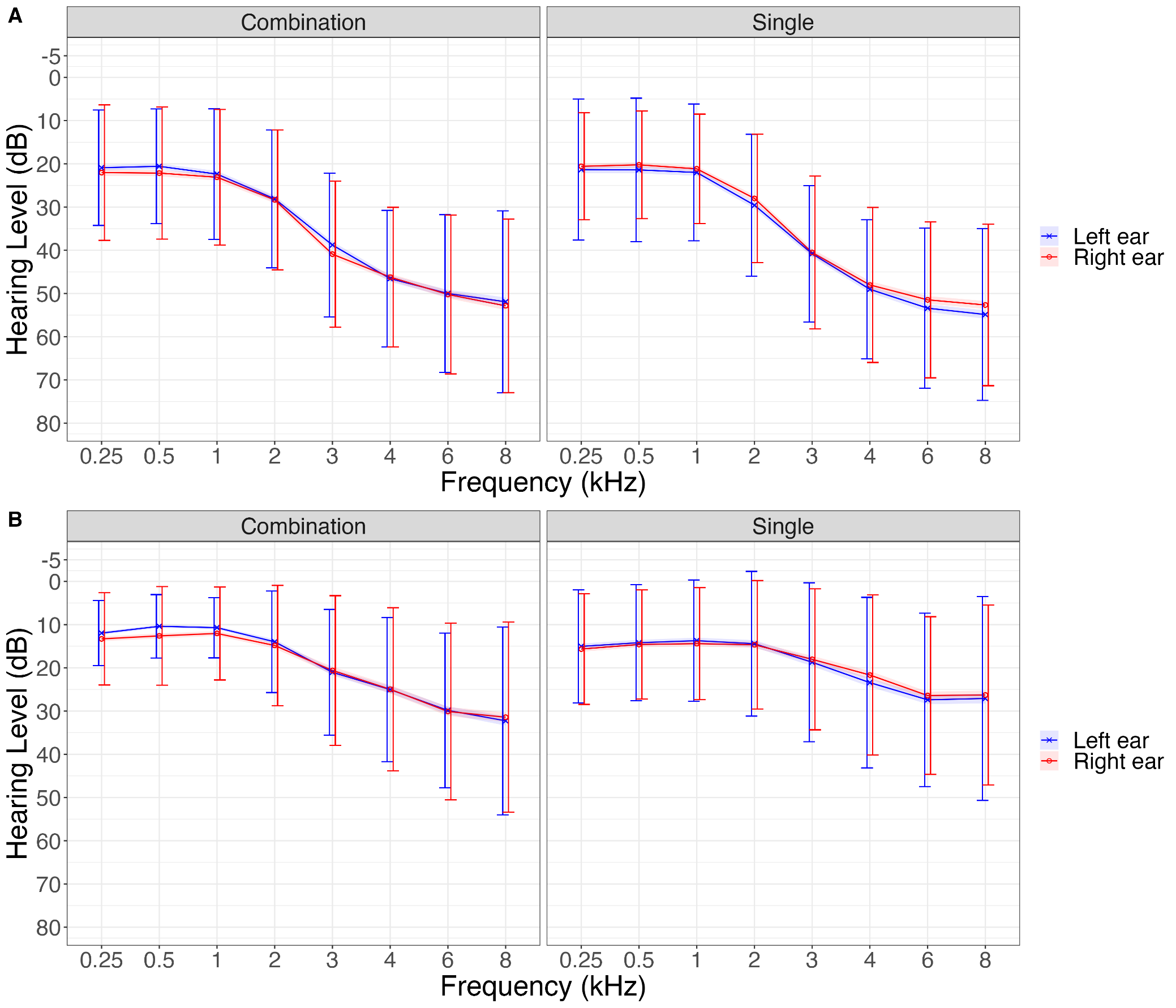

*Note.* Pooled audiogram for patients (A) with hearing aid indication and (B) without hearing aid indication separated for single and combined treatment groups measured by pure tone audiometry at baseline visit according to the guidelines of the British Society of Audiology. Error bars represent standard deviation.

15. ***Figure S4: Audiogram for Patients in all Treatment Groups***

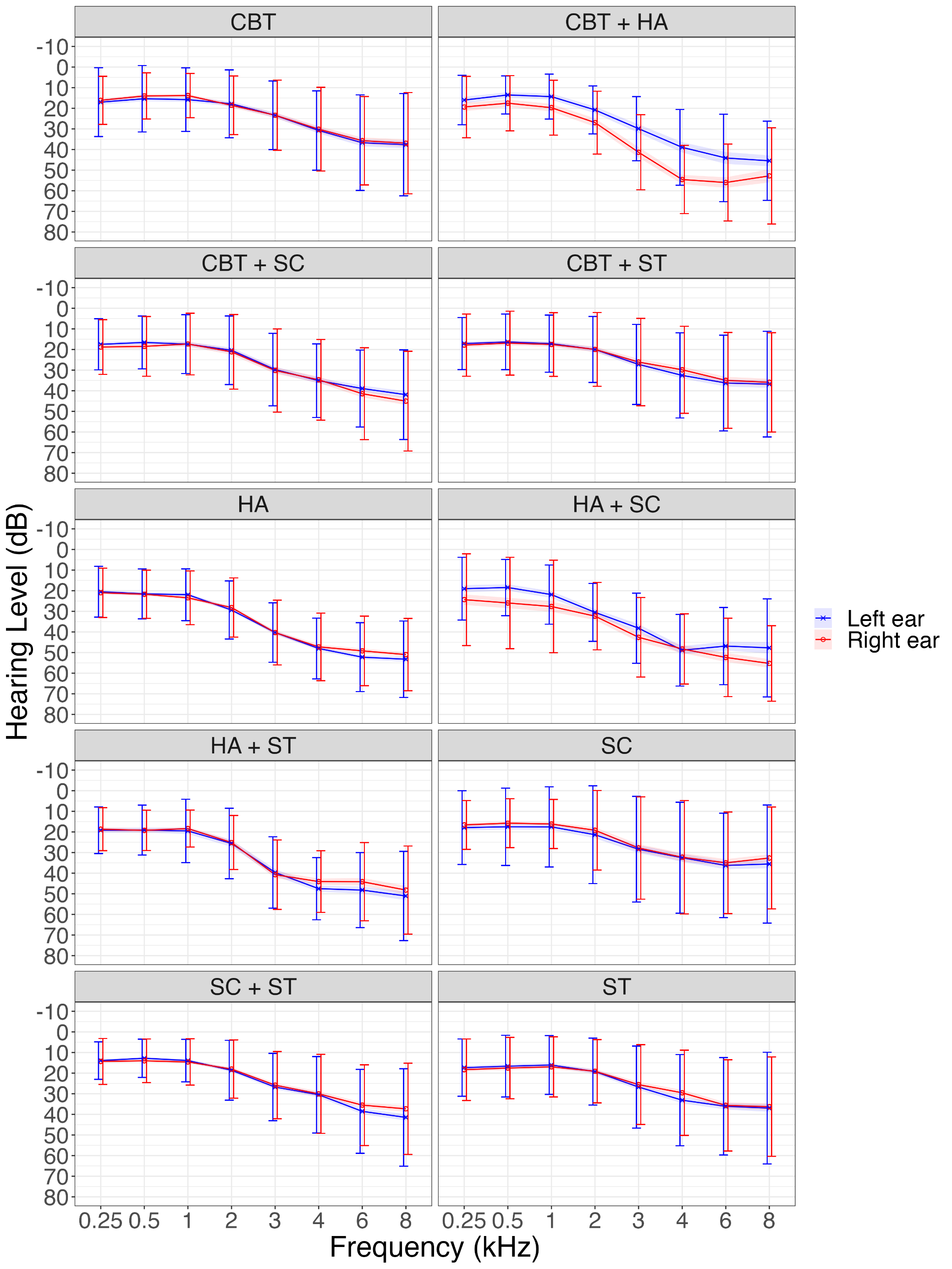

*Note.* Pooled audiograms of patients in all treatment groups measured by pure tone audiometry at baseline visit according to the guidelines of the British Society of Audiology. Error bars represent standard deviation. CBT = Cognitive-Behavioural Therapy; HA = Hearing Aids; SC = Structured Counselling; ST = Sound Therapy.

16. ***Table S12: Primary Outcome (THI) Objective 1: Model parameters***

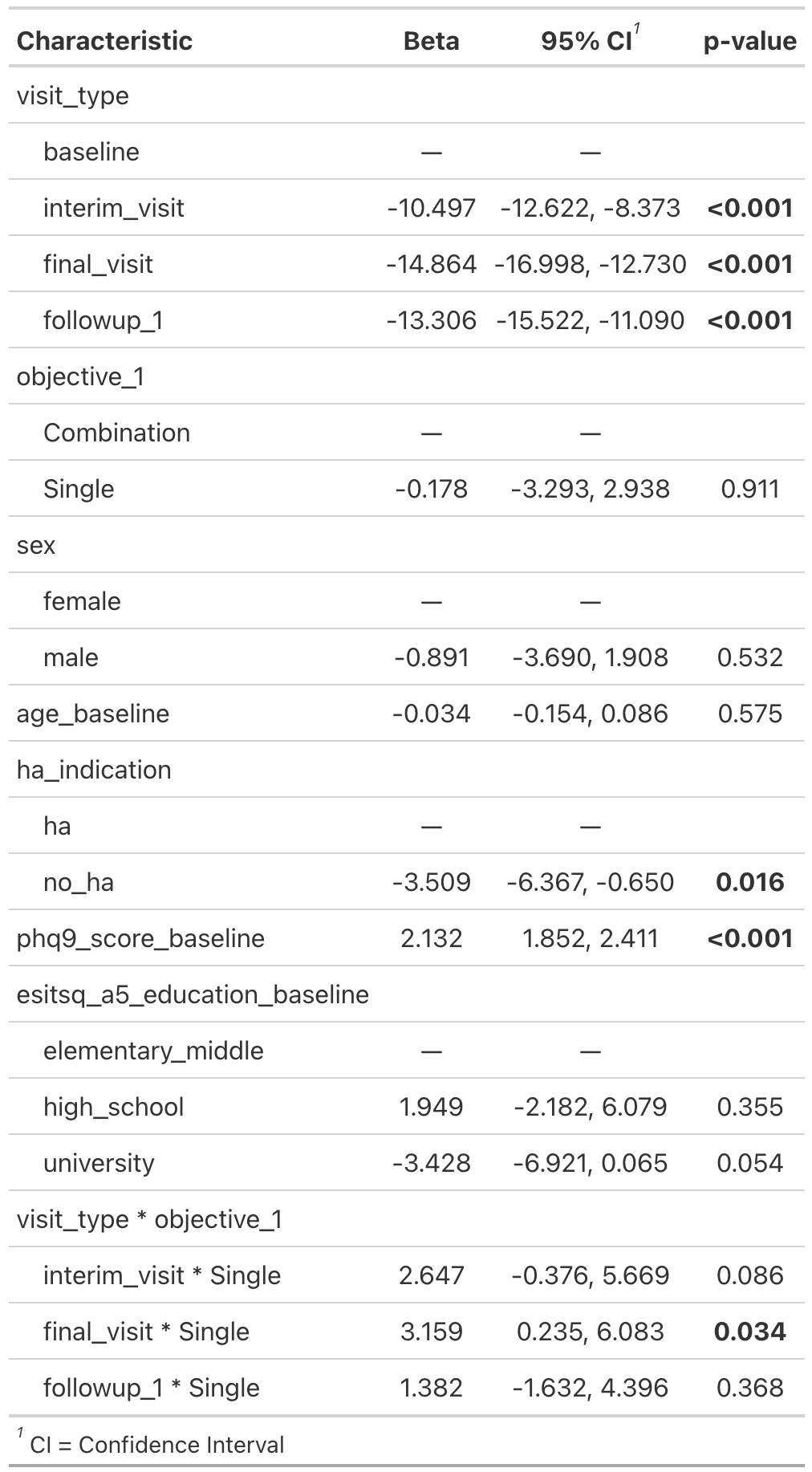

17. ***Table S13: THI change from baseline to final visit for hearing aid indication (yes/no)***

| **Contrast** | **Change from baseline (Hearing aid indication: yes)** | **Change from baseline (Hearing aid indication: no)** |
| --- | --- | --- |
| CBT | -10·4 [-20·9, 0] | -20·2 [-27·1, -13·2] |
| CBT + HA | -15·2 [-26, -4·5] | NA |
| CBT + SC | -18·6 [-28·3, -8·9] | -16·7 [-25·1, -8·4] |
| CBT + ST | -11·4 [-21·3, -1·6] | -15·3 [-22·4, -8·2] |
| HA | -14·4 [-19·6, -9·2] | NA |
| HA + SC | -20 [-29·2, -10·9] | NA |
| HA + ST | -12·9 [-20·5, -5·4] | NA |
| SC | -12·8 [-22·5, -3·1] | -11·6 [-18·3, -4·8] |
| SC + ST | -13·1 [-22·4, -3·9] | -12·5 [-18·7, -6·3] |
| ST | -3 [-12, 6·1] | -4·3 [-10·9, 2·4] |

*Note.* Values depict least-squares mean changes with 95% CI in square brackets. CBT = Cognitive-Behavioural Therapy; HA = Hearing Aids; SC = Structured Counselling; ST = Sound Therapy.

18. ***Table S14: THI change from baseline to final visit for high and low tinnitus distress severity***

| **Contrast** | **Change from baseline (High tinnitus distress severity)** | **Change from baseline (Low tinnitus distress severity)** |
| --- | --- | --- |
| CBT | -25 [-34·2, -15·8] | -9·3 [-16, -2·5] |
| CBT + HA | -18·7 [-36·6, -0·8] | -12·2 [-24, -0·3] |
| CBT + SC | -24·4 [-33·7, -15·1] | -10·6 [-18·6, -2·6] |
| CBT + ST | -23·2 [-32·2, -14·3] | -6·2 [-13·1, 0·7] |
| HA | -18·1 [-26·1, -10·1] | -10·6 [-17·2, -4·1] |
| HA + SC | -26·9 [-41·6, -12·2] | -13·8 [-24·5, -3·2] |
| HA + ST | -12·8 [-24·4, -1·2] | -13·1 [-22·4, -3·8] |
| SC | -15·6 [-24·6, -6·7] | -8·8 [-15·2, -2·5] |
| SC + ST | -19·1 [-27·8, -10·5] | -7·8 [-13·5, -2·1] |
| ST | -6·3 [-14·6, 2·1] | -1·5 [-7·8, 4·8] |

*Note.* Values depict least-squares mean changes with 95% CI in square brackets. High tinnitus distress severity: THI ≥ 48 at Screening. Low tinnitus distress severity: THI < 48 at Screening. CBT = Cognitive-Behavioural Therapy; HA = Hearing Aids; SC = Structured Counselling; ST = Sound Therapy.

19. ***Figure S5: Primary Outcome (THI) Objective 1: Model Assumptions***

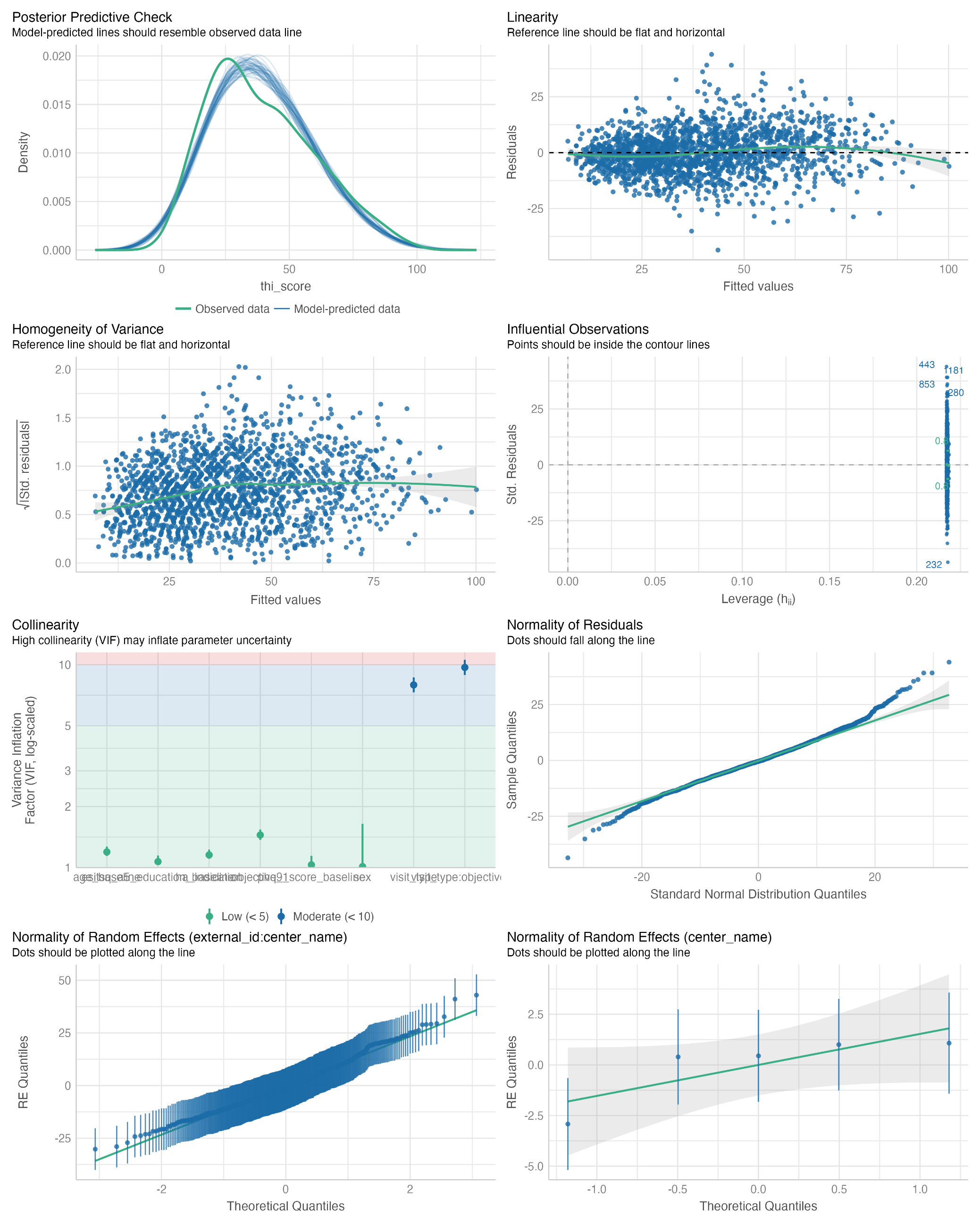

*Note.* Model assumptions are plotted for the first of the 50 imputed datasets.

20. ***Figure S6: Primary Outcome (THI) Objective 2***

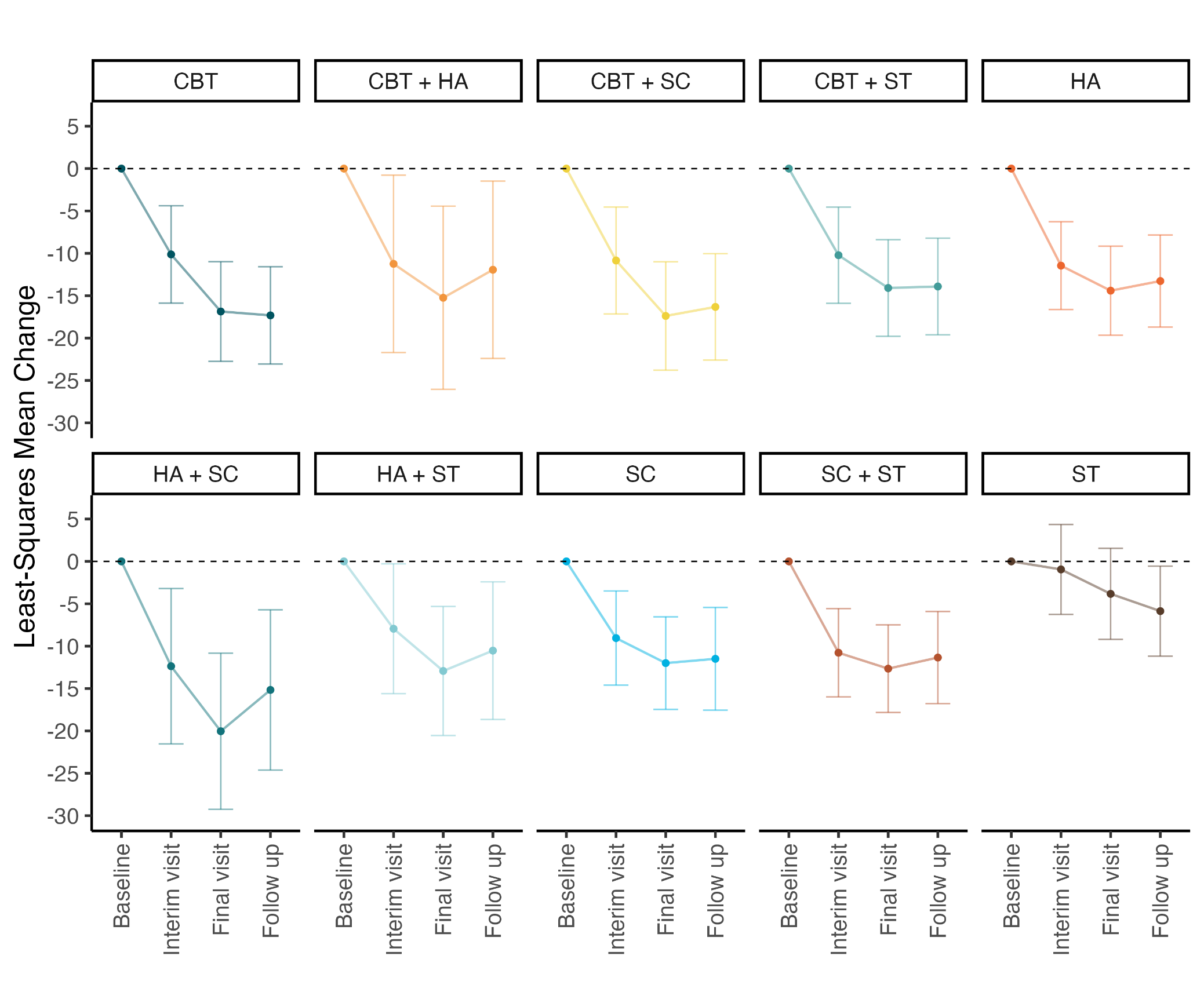

*Note.* Total THI scores range from 0 to 100, with higher scores indicating greater severity of tinnitus. Error bars represent 95% confidence intervals. Sample sizes: CBT: N = 56, HA: N = 59, SC: N = 56, ST: N = 59, CBT + HA: N = 17, CBT + SC: N = 51, CBT + ST: N = 54, HA + SC: N = 19, HA + ST: N = 27, SC + ST: N = 63. CBT = Cognitive-Behavioural Therapy; HA = Hearing Aids; SC = Structured Counselling; ST = Sound Therapy.

21. ***Table S15: Primary Outcome (THI) Objective 2: Post-hoc Comparisons (Baseline to Final Visit)***

| **Contrast (baseline – final visit)** | **Estimate** | **95% CI** | **p** | **p-adjusted** |
| --- | --- | --- | --- | --- |
| CBT - (CBT + HA) | 1·62 | [-7·47, 10·71] | ·727 | 1·00 |
| CBT - (CBT + SC) | -0·53 | [-7·01, 5·96] | ·874 | 1·00 |
| CBT - (CBT + ST) | 2·77 | [-3·56, 9·11] | ·391 | 1·00 |
| CBT - HA | 2·46 | [-3·50, 8·42] | ·419 | 1·00 |
| CBT - (HA + SC) | -3·18 | [-11·50, 5·15] | ·454 | 1·00 |
| CBT - (HA + ST) | 3·93 | [-3·43, 11·29] | ·295 | 1·00 |
| CBT - SC | 4·86 | [-1·26, 10·98] | ·119 | 1·00 |
| CBT - (SC + ST) | 4·21 | [-1·70, 10·11] | ·162 | 1·00 |
| CBT - ST | 13·03 | [6·85, 19·21] | < ·001*** | < ·001*** |
| (CBT + HA) - (CBT + SC) | -2·14 | [-11·60, 7·31] | ·656 | 1·00 |
| (CBT + HA) - (CBT + ST) | 1·16 | [-8·30, 10·61] | ·811 | 1·00 |
| (CBT + HA) - HA | 0·84 | [-8·24, 9·92] | ·856 | 1·00 |
| (CBT + HA) - (HA + SC) | -4·80 | [-15·57, 5·98] | ·383 | 1·00 |
| (CBT + HA) - (HA + ST) | 2·31 | [-7·81, 12·43] | ·654 | 1·00 |
| (CBT + HA) - SC | 3·24 | [-5·97, 12·45] | ·490 | 1·00 |
| (CBT + HA) - (SC + ST) | 2·59 | [-6·56, 11·74] | ·579 | 1·00 |
| (CBT + HA) - ST | 11·41 | [2·21, 20·62] | ·015* | ·675 |
| (CBT + SC) - (CBT + ST) | 3·30 | [-3·38, 9·98] | ·333 | 1·00 |
| (CBT + SC) - HA | 2·98 | [-3·30, 9·26] | ·352 | 1·00 |
| (CBT + SC) - (HA + SC) | -2·65 | [-11·13, 5·83] | ·540 | 1·00 |
| (CBT + SC) - (HA + ST) | 4·46 | [-3·02, 11·93] | ·242 | 1·00 |
| (CBT + SC) - SC | 5·39 | [-1·05, 11·83] | ·101 | 1·00 |
| (CBT + SC) - (SC + ST) | 4·73 | [-1·43, 10·90] | ·132 | 1·00 |
| (CBT + SC) - ST | 13·56 | [7·21, 19·90] | < ·001*** | < ·001*** |
| (CBT + ST) - HA | -0·32 | [-6·22, 5·59] | ·916 | 1·00 |
| (CBT + ST) - (HA + SC) | -5·95 | [-14·30, 2·40] | ·162 | 1·00 |
| (CBT + ST) - (HA + ST) | 1·16 | [-6·07, 8·38] | ·753 | 1·00 |
| (CBT + ST) - SC | 2·09 | [-3·94, 8·11] | ·497 | 1·00 |
| (CBT + ST) - (SC + ST) | 1·43 | [-4·47, 7·34] | ·634 | 1·00 |
| (CBT + ST) - ST | 10·26 | [4·32, 16·19] | ·001** | ·045* |
| HA - (HA + SC) | -5·63 | [-13·75, 2·48] | ·173 | 1·00 |
| HA - (HA + ST) | 1·48 | [-5·56, 8·51] | ·681 | 1·00 |
| HA - SC | 2·40 | [-3·36, 8·17] | ·413 | 1·00 |
| HA - (SC + ST) | 1·75 | [-3·84, 7·34] | ·539 | 1·00 |
| HA - ST | 10·57 | [4·78, 16·37] | < ·001*** | < ·001*** |
| (HA + SC) - (HA + ST) | 7·11 | [-2·05, 16·27] | ·128 | 1·00 |
| (HA + SC) - SC | 8·04 | [-0·09, 16·16] | ·053 | 1·00 |
| (HA + SC) - (SC + ST) | 7·38 | [-0·65, 15·42] | ·072 | 1·00 |
| (HA + SC) - ST | 16·21 | [8·06, 24·36] | < ·001*** | < ·001*** |
| (HA + ST) - SC | 0·93 | [-6·23, 8·09] | ·799 | 1·00 |
| (HA + ST) - (SC + ST) | 0·28 | [-6·75, 7·30] | ·939 | 1·00 |
| (HA + ST) - ST | 9·10 | [2·03, 16·16] | ·012* | ·540 |
| SC - (SC + ST) | -0·65 | [-6·39, 5·08] | ·823 | 1·00 |
| SC - ST | 8·17 | [2·29, 14·05] | ·006** | ·270 |
| (SC + ST) - ST | 8·82 | [3·20, 14·45] | ·002** | ·090 |

*Note.* Objective 2: Pairwise comparison of treatments for the THI difference from baseline to final visit (post-hoc tests). P-values were adjusted for multiple comparisons using the Bonferroni method. CBT = Cognitive-Behavioural Therapy; HA = Hearing Aids; SC = Structured Counselling; ST = Sound Therapy. *p<0·05, **p<0·01, ***p<0·001.

22. ***Table S16: Primary Outcome (THI) Objective 4 & 5 at Final Visit***

| **Objective** | **Contrast** | **Change from baseline** | **95% CI** |
| --- | --- | --- | --- |
| 4_A | Doesnt_have_CBT | -11·57 | [-14·01, -9·14] |
| 4_A | Has_CBT | -16·02 | [-19·33, -12·70] |
| 4_B | Doesnt_have_HA | -11·42 | [-15·47, -7·38] |
| 4_B | Has_HA | -15·07 | [-18·75, -11·39] |
| 4_C | Doesnt_have_SC | -12·46 | [-15·01, -9·91] |
| 4_C | Has_SC | -14·48 | [-17·62, -11·34] |
| 4_D | Doesnt_have_ST | -15·48 | [-18·23, -12·72] |
| 4_D | Has_ST | -10·51 | [-13·43, -7·58] |
| 5 | Brain | -15·36 | [-18·82, -11·89] |
| 5 | Brain_and_Ear | -14·36 | [-17·69, -11·04] |
| 5 | Ear | -9·83 | [-13·20, -6·45] |

*Note.* Values depict least-squares mean changes with 95% CI in square brackets. Objective 4: Treatments with vs. treatments without; Objective 5: Ear mediated vs. brain mediated vs. ear and brain mediated treatments. CBT = Cognitive-Behavioural Therapy; HA = Hearing Aids; SC = Structured Counselling; ST = Sound Therapy.

23. ***Table S17: Primary Outcome (THI) at Interim Visit***

| **Objective** | **Contrast** | **Change from baseline** | **95% CI** |
| --- | --- | --- | --- |
| 1 | Combination | -10·50 | [-13·28, -7·71] |
| 1 | Single | -7·85 | [-10·66, -5·04] |
| 2 | CBT | -10·13 | [-15·87, -4·38] |
| 2 | CBT + HA | -11·24 | [-21·70, -0·78] |
| 2 | CBT + SC | -10·84 | [-17·15, -4·54] |
| 2 | CBT + ST | -10·22 | [-15·90, -4·55] |
| 2 | HA | -11·45 | [-16·63, -6·27] |
| 2 | HA + SC | -12·37 | [-21·53, -3·21] |
| 2 | HA + ST | -7·95 | [-15·61, -0·29] |
| 2 | SC | -9·05 | [-14·60, -3·50] |
| 2 | SC + ST | -10·78 | [-15·99, -5·57] |
| 2 | ST | -0·95 | [-6·25, 4·35] |
| 3_A | Combi_CBT | -10·62 | [-14·57, -6·68] |
| 3_A | Single_CBT | -10·13 | [-15·92, -4·33] |
| 3_B | Combi_HA | -10·17 | [-15·05, -5·28] |
| 3_B | Single_HA | -11·45 | [-16·42, -6·48] |
| 3_C | Combi_SC | -11·03 | [-14·72, -7·34] |
| 3_C | Single_SC | -9·05 | [-14·60, -3·50] |
| 3_D | Combi_ST | -10·04 | [-13·57, -6·51] |
| 3_D | Single_ST | -0·95 | [-6·35, 4·46] |
| 4_A | Doesnt_have_CBT | -8·36 | [-10·83, -5·90] |
| 4_A | Has_CBT | -10·47 | [-13·70, -7·23] |
| 4_B | Doesnt_have_HA | -7·77 | [-11·79, -3·76] |
| 4_B | Has_HA | -10·79 | [-14·39, -7·19] |
| 4_C | Doesnt_have_SC | -8·30 | [-10·87, -5·72] |
| 4_C | Has_SC | -10·44 | [-13·54, -7·35] |
| 4_D | Doesnt_have_ST | -10·58 | [-13·23, -7·92] |
| 4_D | Has_ST | -7·40 | [-10·34, -4·45] |
| 5 | Brain | -9·98 | [-13·33, -6·63] |
| 5 | Brain_and_Ear | -10·83 | [-14·19, -7·47] |
| 5 | Ear | -6·53 | [-9·91, -3·14] |

*Note.* Values depict least-squares mean changes with 95% CI in square brackets. Objective 1: Single vs. Combination (all treatments); Objective 2: All treatments against each other; Objective 3: Single vs. Combination (separately); Objective 4: Treatments with vs. treatments without; Objective 5: Ear-mediated vs. brain-mediated vs. ear- and brain-mediated treatments. CBT = Cognitive-Behavioural Therapy; HA = Hearing Aids; SC = Structured Counselling; ST = Sound Therapy.

24. ***Table S18: Primary Outcome (THI) at Follow-up***

| **Objective** | **Contrast** | **Change from baseline** | **95% CI** |
| --- | --- | --- | --- |
| 1 | Combination | -13·31 | [-16·21, -10·40] |
| 1 | Single | -11·92 | [-14·75, -9·09] |
| 2 | CBT | -17·32 | [-23·06, -11·58] |
| 2 | CBT + HA | -11·94 | [-22·40, -1·48] |
| 2 | CBT + SC | -16·32 | [-22·60, -10·04] |
| 2 | CBT + ST | -13·91 | [-19·62, -8·20] |
| 2 | HA | -13·27 | [-18·70, -7·83] |
| 2 | HA + SC | -15·17 | [-24·62, -5·71] |
| 2 | HA + ST | -10·53 | [-18·65, -2·41] |
| 2 | SC | -11·50 | [-17·55, -5·44] |
| 2 | SC + ST | -11·35 | [-16·77, -5·92] |
| 2 | ST | -5·87 | [-11·18, -0·55] |
| 3_A | Combi_CBT | -14·64 | [-18·56, -10·72] |
| 3_A | Single_CBT | -17·32 | [-23·11, -11·53] |
| 3_B | Combi_HA | -12·31 | [-17·40, -7·21] |
| 3_B | Single_HA | -13·27 | [-18·51, -8·03] |
| 3_C | Combi_SC | -13·80 | [-17·67, -9·93] |
| 3_C | Single_SC | -11·50 | [-17·55, -5·44] |
| 3_D | Combi_ST | -12·15 | [-15·79, -8·52] |
| 3_D | Single_ST | -5·87 | [-11·28, -0·45] |
| 4_A | Doesnt_have_CBT | -10·81 | [-13·39, -8·23] |
| 4_A | Has_CBT | -15·49 | [-18·75, -12·22] |
| 4_B | Doesnt_have_HA | -11·46 | [-15·62, -7·29] |
| 4_B | Has_HA | -12·77 | [-16·59, -8·95] |
| 4_C | Doesnt_have_SC | -12·27 | [-14·90, -9·64] |
| 4_C | Has_SC | -13·12 | [-16·48, -9·75] |
| 4_D | Doesnt_have_ST | -14·42 | [-17·15, -11·69] |
| 4_D | Has_ST | -10·33 | [-13·30, -7·36] |
| 5 | Brain | -15·01 | [-18·56, -11·45] |
| 5 | Brain_and_Ear | -12·79 | [-16·23, -9·35] |
| 5 | Ear | -9·75 | [-13·22, -6·28] |

*Note.* Values depict least-squares mean changes with 95% CI in square brackets. Objective 1: Single vs. Combination (all treatments); Objective 2: All treatments against each other; Objective 3: Single vs. Combination (separately); Objective 4: Treatments with vs. treatments without; Objective 5: Ear-mediated vs. brain-mediated vs. ear- and brain-mediated treatments. CBT = Cognitive-Behavioural Therapy; HA = Hearing Aids; SC = Structured Counselling; ST = Sound Therapy.

25. ***Table S19: Secondary Outcome (TFI) Objective 4 & 5 at Final Visit***

| **Objective** | **Contrast** | **Change from baseline** | **95% CI** |
| --- | --- | --- | --- |
| 4_A | Doesnt_have_CBT | -10·01 | [-12·66, -7·36] |
| 4_A | Has_CBT | -13·33 | [-16·89, -9·78] |
| 4_B | Doesnt_have_HA | -9·25 | [-13·64, -4·86] |
| 4_B | Has_HA | -14·19 | [-18·09, -10·29] |
| 4_C | Doesnt_have_SC | -12·21 | [-14·96, -9·46] |
| 4_C | Has_SC | -9·97 | [-13·33, -6·61] |
| 4_D | Doesnt_have_ST | -12·80 | [-15·62, -9·99] |
| 4_D | Has_ST | -9·37 | [-12·56, -6·18] |
| 5 | Brain | -12·26 | [-15·89, -8·64] |
| 5 | Brain_and_Ear | -11·13 | [-14·86, -7·40] |
| 5 | Ear | -10·37 | [-14·00, -6·73] |

*Note.* Values depict least-squares mean changes with 95% CI in square brackets. Objective 4: Treatments with vs. treatments without; Objective 5: Ear-mediated vs. brain-mediated vs. ear- and brain-mediated treatments. CBT = Cognitive-Behavioural Therapy; HA = Hearing Aids; SC = Structured Counselling; ST = Sound Therapy.

26. ***Table S20: Secondary Outcome (Mini-TQ) Objective 4 & 5 at Final Visit***

| **Objective** | **Contrast** | **Change from baseline** | **95% CI** |
| --- | --- | --- | --- |
| 4_A | Doesnt_have_CBT | -2·63 | [-3·25, -2·02] |
| 4_A | Has_CBT | -3·91 | [-4·71, -3·10] |
| 4_B | Doesnt_have_HA | -2·51 | [-3·49, -1·53] |
| 4_B | Has_HA | -3·28 | [-4·16, -2·41] |
| 4_C | Doesnt_have_SC | -3·05 | [-3·69, -2·41] |
| 4_C | Has_SC | -3·23 | [-3·99, -2·48] |
| 4_D | Doesnt_have_ST | -3·61 | [-4·27, -2·95] |
| 4_D | Has_ST | -2·50 | [-3·24, -1·77] |
| 5 | Brain | -3·65 | [-4·50, -2·80] |
| 5 | Brain_and_Ear | -3·30 | [-4·15, -2·44] |
| 5 | Ear | -2·35 | [-3·21, -1·50] |

*Note.* Values depict least-squares mean changes with 95% CI in square brackets. Objective 4: Treatments with vs. treatments without; Objective 5: Ear-mediated vs. brain-mediated vs. ear- and brain-mediated treatments. CBT = Cognitive-Behavioural Therapy; HA = Hearing Aids; SC = Structured Counselling; ST = Sound Therapy.

27. ***Table S21: Secondary Outcome (NRS-2: “How strong or loud is your tinnitus at present?”) Objective 4 & 5 at Final Visit***

| **Objective** | **Contrast** | **Change from baseline** | **95% CI** |
| --- | --- | --- | --- |
| 4_A | Doesnt_have_CBT | -0·80 | [-1·15, -0·45] |
| 4_A | Has_CBT | -0·73 | [-1·21, -0·25] |
| 4_B | Doesnt_have_HA | -0·44 | [-1·03, 0·15] |
| 4_B | Has_HA | -1·10 | [-1·63, -0·57] |
| 4_C | Doesnt_have_SC | -0·79 | [-1·15, -0·42] |
| 4_C | Has_SC | -0·76 | [-1·20, -0·31] |
| 4_D | Doesnt_have_ST | -0·87 | [-1·27, -0·48] |
| 4_D | Has_ST | -0·64 | [-1·05, -0·24] |
| 5 | Brain | -0·75 | [-1·26, -0·24] |
| 5 | Brain_and_Ear | -0·68 | [-1·17, -0·19] |
| 5 | Ear | -0·90 | [-1·38, -0·42] |

*Note.* Values depict least-squares mean changes with 95% CI in square brackets. Objective 4: Treatments with vs. treatments without; Objective 5: Ear-mediated vs. brain-mediated vs. ear- and brain-mediated treatments. CBT = Cognitive-Behavioural Therapy; HA = Hearing Aids; SC = Structured Counselling; ST = Sound Therapy.

28. ***Table S22: Secondary Outcome (PHQ-9) Objective 4 & 5 at Final Visit***

| **Objective** | **Contrast** | **Change from baseline** | **95% CI** |
| --- | --- | --- | --- |
| 4_A | Doesnt_have_CBT | -1·46 | [-2·04, -0·88] |
| 4_A | Has_CBT | -1·70 | [-2·45, -0·95] |
| 4_B | Doesnt_have_HA | -1·28 | [-2·25, -0·32] |
| 4_B | Has_HA | -1·90 | [-2·76, -1·05] |
| 4_C | Doesnt_have_SC | -1·62 | [-2·22, -1·02] |
| 4_C | Has_SC | -1·46 | [-2·20, -0·73] |
| 4_D | Doesnt_have_ST | -1·85 | [-2·48, -1·23] |
| 4_D | Has_ST | -1·17 | [-1·87, -0·48] |
| 5 | Brain | -1·72 | [-2·52, -0·93] |
| 5 | Brain_and_Ear | -1·36 | [-2·17, -0·55] |
| 5 | Ear | -1·57 | [-2·37, -0·77] |

*Note.* Values depict least-squares mean changes with 95% CI in square brackets. Objective 4: Treatments with vs. treatments without; Objective 5: Ear-mediated vs. brain-mediated vs. ear- and brain-mediated treatments. CBT = Cognitive-Behavioural Therapy; HA = Hearing Aids; SC = Structured Counselling; ST = Sound Therapy.

29. ***Figure S7: Secondary Outcome (CGI-I) at Final Visit – Single vs. Combination (Objective 1)***

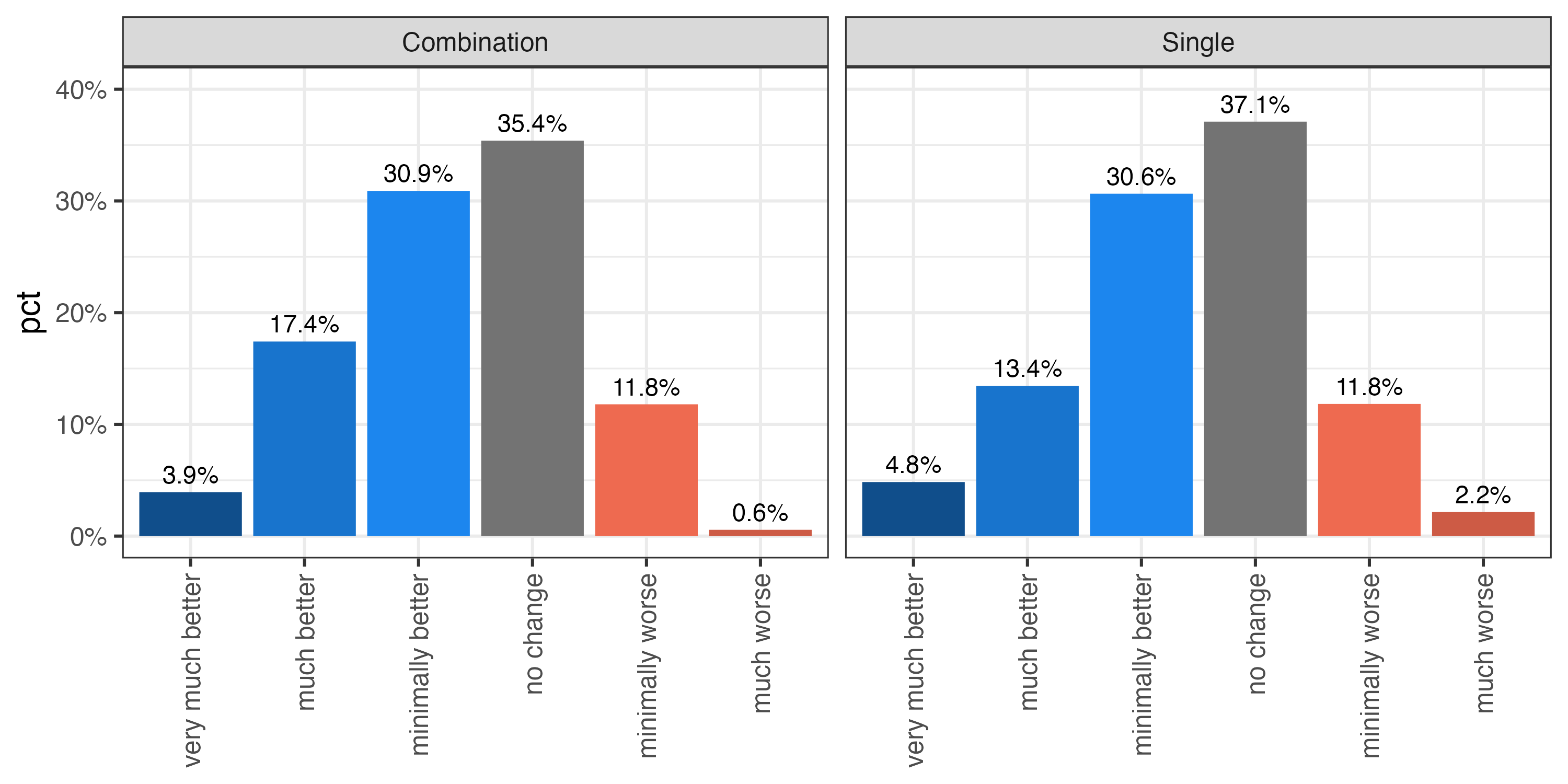

*Note.* Distribution (in percentage) of CGI-I (Clinical Global Impression Scale) at final visit for Objective 1.

30. ***Figure S8: Secondary Outcome (CGI-I) at Final Visit – all Treatment Arms (Objective 2)***

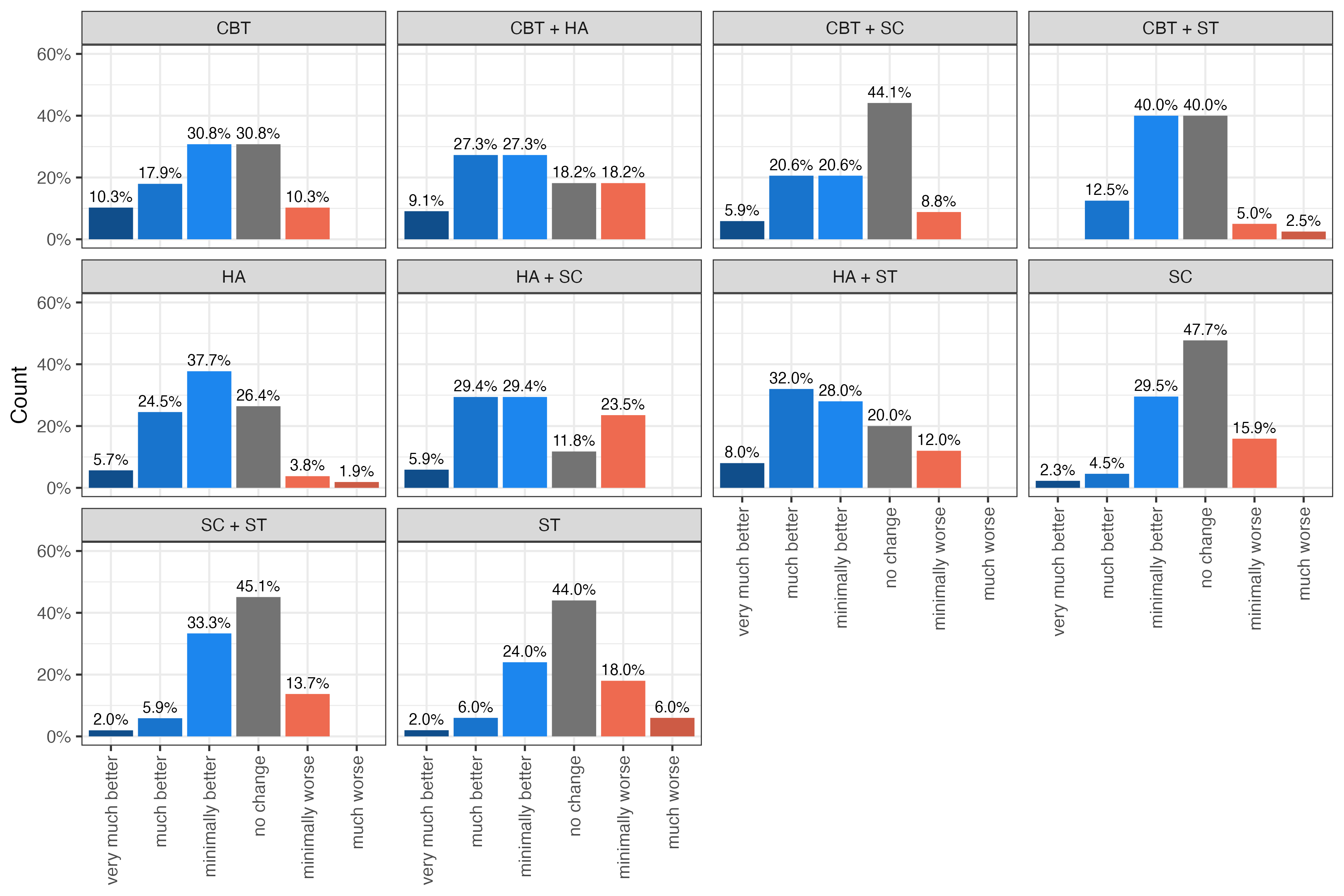

*Note.* Distribution (in percentage) of CGI-I (Clinical Global Impression Scale) at final visit for Objective 2. CBT = Cognitive-Behavioural Therapy; HA = Hearing Aids; SC = Structured Counselling; ST = Sound Therapy.

31. ***Figure S9: Secondary Outcome (CGI-I) at Final Visit – all Treatment Arms (Objective 2) in Patients With and Without Hearing Aid Indication***

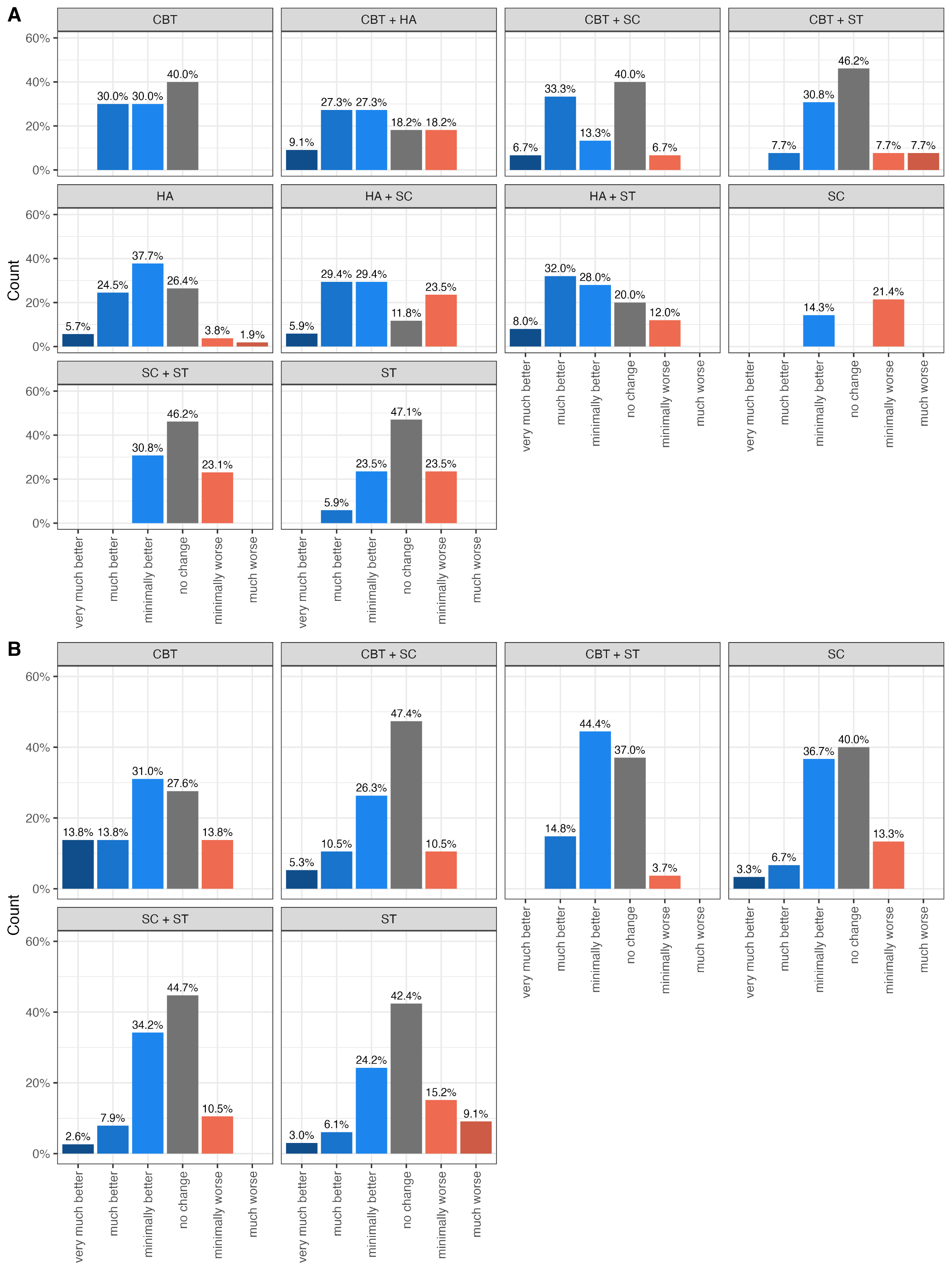

*Note.* Distribution (in percentage) of CGI-I (Clinical Global Impression Scale) at final visit for Objective 2 in patients (A) with hearing aid indication and (B) without hearing aid indication. CBT = Cognitive-Behavioural Therapy; HA = Hearing Aids; SC = Structured Counselling; ST = Sound Therapy.

32. ***Figure S10: Secondary Outcome (CGI-I) at Final Visit – all Treatment Arms (Objective 2) in Patients with High and Low Tinnitus Distress Severity***

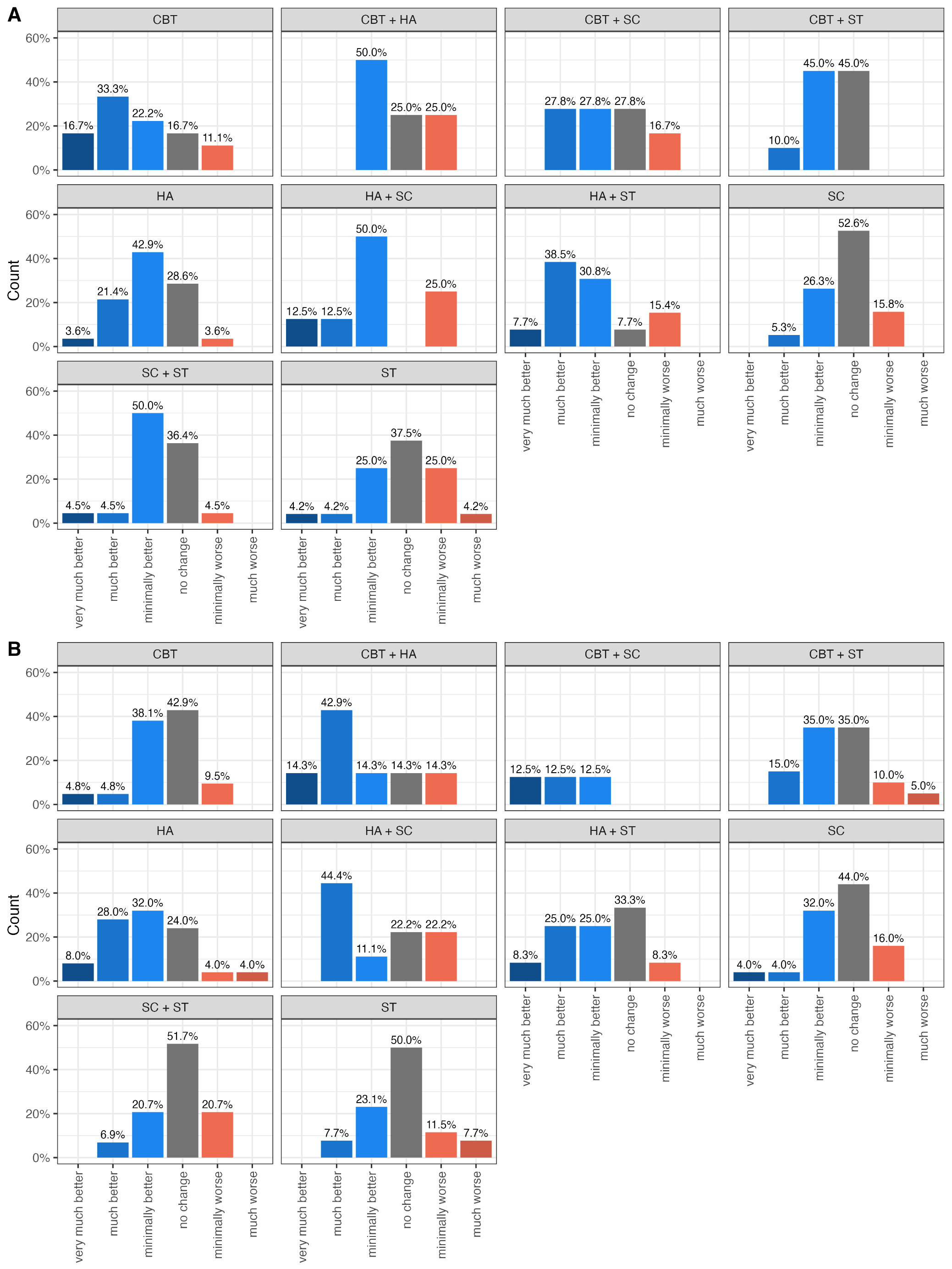

*Note.* Distribution (in percentage) of CGI-I (Clinical Global Impression Scale) at final visit for Objective 2 in patients (A) with High and (B) Low tinnitus distress severity. High tinnitus distress severity: THI ≥ 48 at Screening. Low tinnitus distress severity: THI < 48 at Screening. CBT = Cognitive-Behavioural Therapy; HA = Hearing Aids; SC = Structured Counselling; ST = Sound Therapy.

33. ***Table S23: Secondary Outcome (WHO-QoL-Bref 1: Physical Health) at Final***

***Visit***

| **Objective** | **Contrast** | **Change from baseline** | **95% CI** |
| --- | --- | --- | --- |
| 1 | Combination | 0·29 | [0·01, 0·58] |
| 1 | Single | 0·42 | [0·13, 0·71] |
| 2 | CBT | 0·43 | [-0·18, 1·04] |
| 2 | CBT + HA | 0·49 | [-0·64, 1·62] |
| 2 | CBT + SC | 0·24 | [-0·39, 0·88] |
| 2 | CBT + ST | 0·36 | [-0·23, 0·95] |
| 2 | HA | 0·52 | [-0·01, 1·06] |
| 2 | HA + SC | 0·64 | [-0·32, 1·60] |
| 2 | HA + ST | 0·36 | [-0·43, 1·15] |
| 2 | SC | 0·20 | [-0·39, 0·79] |
| 2 | SC + ST | 0·10 | [-0·44, 0·63] |
| 2 | ST | 0·51 | [-0·04, 1·07] |
| 3_A | Combi_CBT | 0·33 | [-0·07, 0·73] |
| 3_A | Single_CBT | 0·43 | [-0·17, 1·03] |
| 3_B | Combi_HA | 0·48 | [-0·06, 1·01] |
| 3_B | Single_HA | 0·52 | [-0·01, 1·06] |
| 3_C | Combi_SC | 0·23 | [-0·15, 0·60] |
| 3_C | Single_SC | 0·20 | [-0·38, 0·79] |
| 3_D | Combi_ST | 0·24 | [-0·12, 0·61] |
| 3_D | Single_ST | 0·51 | [-0·05, 1·07] |
| 4_A | Doesnt_have_CBT | 0·35 | [0·10, 0·61] |
| 4_A | Has_CBT | 0·36 | [0·02, 0·70] |
| 4_B | Doesnt_have_HA | 0·34 | [-0·08, 0·75] |
| 4_B | Has_HA | 0·50 | [0·13, 0·87] |
| 4_C | Doesnt_have_SC | 0·45 | [0·18, 0·72] |
| 4_C | Has_SC | 0·22 | [-0·10, 0·54] |
| 4_D | Doesnt_have_ST | 0·38 | [0·11, 0·66] |
| 4_D | Has_ST | 0·32 | [0·02, 0·62] |
| 5 | Brain | 0·29 | [-0·06, 0·64] |
| 5 | Brain_and_Ear | 0·30 | [-0·05, 0·65] |
| 5 | Ear | 0·49 | [0·14, 0·84] |

*Note.* Values depict least-squares mean changes with 95% CI in square brackets. Objective 1: Single vs. Combination (all treatments); Objective 2: All treatments against each other; Objective 3: Single vs. Combination (separately); Objective 4: Treatments with vs. treatments without; Objective 5: Ear-mediated vs. brain-mediated vs. ear- and brain-mediated treatments. CBT = Cognitive-Behavioural Therapy; HA = Hearing Aids; SC = Structured Counselling; ST = Sound Therapy.

34. ***Table S24: Secondary Outcome (WHO-QoL-Bref 2: Psychological Health) at Final Visit***

| **Objective** | **Contrast** | **Change from baseline** | **95% CI** |
| --- | --- | --- | --- |
| 1 | Combination | 0·36 | [0·07, 0·65] |
| 1 | Single | 0·36 | [0·08, 0·65] |
| 2 | CBT | 0·33 | [-0·25, 0·92] |
| 2 | CBT + HA | 0·75 | [-0·35, 1·86] |
| 2 | CBT + SC | 0·28 | [-0·35, 0·91] |
| 2 | CBT + ST | 0·35 | [-0·27, 0·97] |
| 2 | HA | 0·35 | [-0·18, 0·89] |
| 2 | HA + SC | 0·15 | [-0·81, 1·10] |
| 2 | HA + ST | 0·41 | [-0·37, 1·20] |
| 2 | SC | 0·30 | [-0·28, 0·88] |
| 2 | SC + ST | 0·35 | [-0·19, 0·89] |
| 2 | ST | 0·47 | [-0·09, 1·02] |
| 3_A | Combi_CBT | 0·38 | [-0·05, 0·81] |
| 3_A | Single_CBT | 0·33 | [-0·26, 0·93] |
| 3_B | Combi_HA | 0·43 | [-0·08, 0·94] |
| 3_B | Single_HA | 0·35 | [-0·16, 0·87] |
| 3_C | Combi_SC | 0·30 | [-0·08, 0·68] |
| 3_C | Single_SC | 0·30 | [-0·29, 0·89] |
| 3_D | Combi_ST | 0·36 | [-0·01, 0·73] |
| 3_D | Single_ST | 0·47 | [-0·10, 1·04] |
| 4_A | Doesnt_have_CBT | 0·36 | [0·11, 0·61] |
| 4_A | Has_CBT | 0·36 | [0·02, 0·70] |
| 4_B | Doesnt_have_HA | 0·34 | [-0·09, 0·76] |
| 4_B | Has_HA | 0·39 | [0·03, 0·75] |
| 4_C | Doesnt_have_SC | 0·40 | [0·14, 0·66] |
| 4_C | Has_SC | 0·30 | [-0·02, 0·61] |
| 4_D | Doesnt_have_ST | 0·34 | [0·06, 0·61] |
| 4_D | Has_ST | 0·39 | [0·09, 0·70] |
| 5 | Brain | 0·31 | [-0·05, 0·66] |
| 5 | Brain_and_Ear | 0·37 | [0·01, 0·73] |
| 5 | Ear | 0·41 | [0·06, 0·75] |

*Note.* Values depict least-squares mean changes with 95% CI in square brackets. Objective 1: Single vs. Combination (all treatments); Objective 2: All treatments against each other; Objective 3: Single vs. Combination (separately); Objective 4: Treatments with vs. treatments without; Objective 5: Ear-mediated vs. brain-mediated vs. ear- and brain-mediated treatments. CBT = Cognitive-Behavioural Therapy; HA = Hearing Aids; SC = Structured Counselling; ST = Sound Therapy.

35. ***Table S25: Secondary Outcome (WHO-QoL-Bref 3: Social Factors) at Final***

***Visit***

| **Objective** | **Contrast** | **Change from baseline** | **95% CI** |
| --- | --- | --- | --- |
| 1 | Combination | 0·17 | [-0·26, 0·60] |
| 1 | Single | 0·20 | [-0·22, 0·63] |
| 2 | CBT | 0·26 | [-0·62, 1·14] |
| 2 | CBT + HA | 0·49 | [-1·14, 2·12] |
| 2 | CBT + SC | 0·62 | [-0·35, 1·59] |
| 2 | CBT + ST | 0·14 | [-0·76, 1·04] |
| 2 | HA | 0·63 | [-0·18, 1·43] |
| 2 | HA + SC | 0·93 | [-0·48, 2·34] |
| 2 | HA + ST | -0·09 | [-1·27, 1·10] |
| 2 | SC | 0·01 | [-0·86, 0·88] |
| 2 | SC + ST | -0·38 | [-1·19, 0·42] |
| 2 | ST | -0·09 | [-0·93, 0·75] |
| 3_A | Combi_CBT | 0·39 | [-0·23, 1·01] |
| 3_A | Single_CBT | 0·26 | [-0·62, 1·14] |
| 3_B | Combi_HA | 0·38 | [-0·40, 1·15] |
| 3_B | Single_HA | 0·63 | [-0·16, 1·41] |
| 3_C | Combi_SC | 0·19 | [-0·35, 0·73] |
| 3_C | Single_SC | 0·01 | [-0·82, 0·84] |
| 3_D | Combi_ST | -0·13 | [-0·69, 0·43] |
| 3_D | Single_ST | -0·09 | [-0·97, 0·79] |
| 4_A | Doesnt_have_CBT | 0·08 | [-0·31, 0·47] |
| 4_A | Has_CBT | 0·35 | [-0·16, 0·86] |
| 4_B | Doesnt_have_HA | 0·30 | [-0·34, 0·94] |
| 4_B | Has_HA | 0·50 | [-0·07, 1·06] |
| 4_C | Doesnt_have_SC | 0·22 | [-0·18, 0·62] |
| 4_C | Has_SC | 0·14 | [-0·35, 0·62] |
| 4_D | Doesnt_have_ST | 0·43 | [0·01, 0·84] |
| 4_D | Has_ST | -0·12 | [-0·58, 0·34] |
| 5 | Brain | 0·29 | [-0·24, 0·81] |
| 5 | Brain_and_Ear | 0·06 | [-0·47, 0·59] |
| 5 | Ear | 0·20 | [-0·32, 0·73] |

*Note.* Values depict least-squares mean changes with 95% CI in square brackets. Objective 1: Single vs. Combination (all treatments); Objective 2: All treatments against each other; Objective 3: Single vs. Combination (separately); Objective 4: Treatments with vs. treatments without; Objective 5: Ear-mediated vs. brain-mediated vs. ear- and brain-mediated treatments. CBT = Cognitive-Behavioural Therapy; HA = Hearing Aids; SC = Structured Counselling; ST = Sound Therapy.

36. ***Table S26: Secondary Outcome (WHO-QoL-Bref 4: Environment) at Final***

***Visit***

| **Objective** | **Contrast** | **Change from baseline** | **95% CI** |
| --- | --- | --- | --- |
| 1 | Combination | 0·28 | [-0·04, 0·61] |
| 1 | Single | 0·35 | [0·02, 0·67] |
| 2 | CBT | 0·36 | [-0·31, 1·02] |
| 2 | CBT + HA | 0·57 | [-0·72, 1·85] |
| 2 | CBT + SC | 0·06 | [-0·64, 0·75] |
| 2 | CBT + ST | 0·46 | [-0·21, 1·13] |
| 2 | HA | 0·74 | [0·14, 1·34] |
| 2 | HA + SC | 0·66 | [-0·44, 1·75] |
| 2 | HA + ST | 0·35 | [-0·53, 1·23] |
| 2 | SC | 0·11 | [-0·54, 0·76] |
| 2 | SC + ST | 0·09 | [-0·52, 0·71] |
| 2 | ST | 0·17 | [-0·47, 0·80] |
| 3_A | Combi_CBT | 0·31 | [-0·13, 0·74] |
| 3_A | Single_CBT | 0·36 | [-0·28, 0·99] |
| 3_B | Combi_HA | 0·50 | [-0·11, 1·11] |
| 3_B | Single_HA | 0·74 | [0·15, 1·33] |
| 3_C | Combi_SC | 0·16 | [-0·27, 0·59] |
| 3_C | Single_SC | 0·11 | [-0·56, 0·78] |
| 3_D | Combi_ST | 0·28 | [-0·13, 0·69] |
| 3_D | Single_ST | 0·17 | [-0·47, 0·81] |
| 4_A | Doesnt_have_CBT | 0·31 | [0·02, 0·60] |
| 4_A | Has_CBT | 0·32 | [-0·06, 0·71] |
| 4_B | Doesnt_have_HA | 0·29 | [-0·20, 0·77] |
| 4_B | Has_HA | 0·62 | [0·21, 1·02] |
| 4_C | Doesnt_have_SC | 0·43 | [0·13, 0·73] |
| 4_C | Has_SC | 0·14 | [-0·21, 0·50] |
| 4_D | Doesnt_have_ST | 0·37 | [0·06, 0·67] |
| 4_D | Has_ST | 0·25 | [-0·10, 0·59] |
| 5 | Brain | 0·18 | [-0·21, 0·57] |
| 5 | Brain_and_Ear | 0·35 | [-0·06, 0·75] |
| 5 | Ear | 0·44 | [0·04, 0·83] |

*Note.* Values depict least-squares mean changes with 95% CI in square brackets. Objective 1: Single vs. Combination (all treatments); Objective 2: All treatments against each other; Objective 3: Single vs. Combination (separately); Objective 4: Treatments with vs. treatments without; Objective 5: Ear-mediated vs. brain-mediated vs. ear- and brain-mediated treatments. CBT = Cognitive-Behavioural Therapy; HA = Hearing Aids; SC = Structured Counselling; ST = Sound Therapy.

37. ***Table S27: Secondary Outcome (NRS 1: “How much of a problem is your tinnitus at present?”) at Final Visit***

| **Objective** | **Contrast** | **Change from baseline** | **95% CI** |
| --- | --- | --- | --- |
| 1 | Combination | -0·53 | [-0·71, -0·35] |
| 1 | Single | -0·49 | [-0·67, -0·31] |
| 2 | CBT | -0·53 | [-0·94, -0·11] |
| 2 | CBT + HA | -0·87 | [-1·55, -0·20] |
| 2 | CBT + SC | -0·54 | [-0·93, -0·14] |
| 2 | CBT + ST | -0·57 | [-0·95, -0·19] |
| 2 | HA | -0·60 | [-0·94, -0·27] |
| 2 | HA + SC | -0·49 | [-1·09, 0·11] |
| 2 | HA + ST | -0·58 | [-1·07, -0·08] |
| 2 | SC | -0·61 | [-0·98, -0·25] |
| 2 | SC + ST | -0·39 | [-0·72, -0·05] |
| 2 | ST | -0·24 | [-0·60, 0·12] |
| 3_A | Combi_CBT | -0·60 | [-0·86, -0·33] |
| 3_A | Single_CBT | -0·53 | [-0·96, -0·09] |
| 3_B | Combi_HA | -0·63 | [-0·95, -0·31] |
| 3_B | Single_HA | -0·60 | [-0·91, -0·29] |
| 3_C | Combi_SC | -0·46 | [-0·69, -0·23] |
| 3_C | Single_SC | -0·61 | [-0·96, -0·27] |
| 3_D | Combi_ST | -0·49 | [-0·72, -0·26] |
| 3_D | Single_ST | -0·24 | [-0·61, 0·13] |
| 4_A | Doesnt_have_CBT | -0·47 | [-0·63, -0·31] |
| 4_A | Has_CBT | -0·58 | [-0·79, -0·36] |
| 4_B | Doesnt_have_HA | -0·38 | [-0·65, -0·12] |
| 4_B | Has_HA | -0·62 | [-0·85, -0·39] |
| 4_C | Doesnt_have_SC | -0·52 | [-0·69, -0·35] |
| 4_C | Has_SC | -0·50 | [-0·71, -0·30] |
| 4_D | Doesnt_have_ST | -0·58 | [-0·75, -0·41] |
| 4_D | Has_ST | -0·42 | [-0·61, -0·23] |
| 5 | Brain | -0·56 | [-0·78, -0·34] |
| 5 | Brain_and_Ear | -0·52 | [-0·74, -0·30] |
| 5 | Ear | -0·45 | [-0·67, -0·23] |

*Note.* Values depict least-squares mean changes with 95% CI in square brackets. Since the outcome is ordinally scaled, it was fitted as a sensitivity analysis with clmm of the ordinal package, as no differences were found to the mixed effect model, the results of the mixed effect model are reported here. Objective 1: Single vs. Combination (all treatments); Objective 2: All treatments against each other; Objective 3: Single vs. Combination (separately); Objective 4: Treatments with vs. treatments without; Objective 5: Ear-mediated vs. brain-mediated vs. ear-and brain-mediated treatments. CBT = Cognitive-Behavioural Therapy; HA = Hearing Aids; SC = Structured Counselling; ST = Sound Therapy.

38. ***Table S28: Secondary Outcome (NRS 3: “How uncomfortable is your tinnitus at present?”) at Final Visit***

| **Objective** | **Contrast** | **Change from baseline** | **95% CI** |
| --- | --- | --- | --- |
| 1 | Combination | -1·68 | [-2·13, -1·23] |
| 1 | Single | -1·25 | [-1·70, -0·80] |
| 2 | CBT | -1·76 | [-2·72, -0·80] |
| 2 | CBT + HA | -1·94 | [-3·71, -0·17] |
| 2 | CBT + SC | -1·94 | [-2·94, -0·93] |
| 2 | CBT + ST | -1·68 | [-2·65, -0·72] |
| 2 | HA | -1·63 | [-2·50, -0·77] |
| 2 | HA + SC | -1·22 | [-2·76, 0·32] |
| 2 | HA + ST | -1·65 | [-2·92, -0·38] |
| 2 | SC | -1·08 | [-2·00, -0·16] |
| 2 | SC + ST | -1·55 | [-2·42, -0·68] |
| 2 | ST | -0·55 | [-1·44, 0·33] |
| 3_A | Combi_CBT | -1·83 | [-2·47, -1·18] |
| 3_A | Single_CBT | -1·76 | [-2·72, -0·80] |
| 3_B | Combi_HA | -1·60 | [-2·49, -0·71] |
| 3_B | Single_HA | -1·63 | [-2·52, -0·75] |
| 3_C | Combi_SC | -1·65 | [-2·25, -1·05] |
| 3_C | Single_SC | -1·08 | [-2·01, -0·15] |
| 3_D | Combi_ST | -1·62 | [-2·20, -1·03] |
| 3_D | Single_ST | -0·55 | [-1·45, 0·34] |
| 4_A | Doesnt_have_CBT | -1·25 | [-1·66, -0·85] |
| 4_A | Has_CBT | -1·80 | [-2·34, -1·27] |
| 4_B | Doesnt_have_HA | -1·14 | [-1·80, -0·47] |
| 4_B | Has_HA | -1·62 | [-2·22, -1·01] |
| 4_C | Doesnt_have_SC | -1·46 | [-1·87, -1·04] |
| 4_C | Has_SC | -1·48 | [-1·98, -0·98] |
| 4_D | Doesnt_have_ST | -1·59 | [-2·02, -1·16] |
| 4_D | Has_ST | -1·31 | [-1·80, -0·82] |
| 5 | Brain | -1·58 | [-2·14, -1·02] |
| 5 | Brain_and_Ear | -1·60 | [-2·16, -1·04] |
| 5 | Ear | -1·20 | [-1·75, -0·64] |

*Note.* Values depict least-squares mean changes with 95% CI in square brackets. Objective 1: Single vs. Combination (all treatments); Objective 2: All treatments against each other; Objective 3: Single vs. Combination (separately); Objective 4: Treatments with vs. treatments without; Objective 5: Ear-mediated vs. brain-mediated vs. ear- and brain-mediated treatments. CBT = Cognitive-Behavioural Therapy; HA = Hearing Aids; SC = Structured Counselling; ST = Sound Therapy.

39. ***Table S29: Secondary outcome (NRS 4: “How annoying is your tinnitus at present?”) at final visit***

| **Objective** | **Contrast** | **Change from baseline** | **95% CI** |
| --- | --- | --- | --- |
| 1 | Combination | -1·53 | [-1·99, -1·08] |
| 1 | Single | -1·18 | [-1·62, -0·73] |
| 2 | CBT | -1·58 | [-2·55, -0·62] |
| 2 | CBT + HA | -2·00 | [-3·73, -0·27] |
| 2 | CBT + SC | -1·83 | [-2·84, -0·81] |
| 2 | CBT + ST | -1·29 | [-2·23, -0·36] |
| 2 | HA | -1·63 | [-2·49, -0·78] |
| 2 | HA + SC | -1·68 | [-3·21, -0·16] |
| 2 | HA + ST | -1·16 | [-2·42, 0·09] |
| 2 | SC | -1·03 | [-1·97, -0·10] |
| 2 | SC + ST | -1·48 | [-2·34, -0·63] |
| 2 | ST | -0·47 | [-1·35, 0·42] |
| 3_A | Combi_CBT | -1·61 | [-2·26, -0·97] |
| 3_A | Single_CBT | -1·58 | [-2·55, -0·62] |
| 3_B | Combi_HA | -1·55 | [-2·43, -0·66] |
| 3_B | Single_HA | -1·63 | [-2·53, -0·73] |
| 3_C | Combi_SC | -1·64 | [-2·24, -1·04] |
| 3_C | Single_SC | -1·03 | [-1·96, -0·10] |
| 3_D | Combi_ST | -1·35 | [-1·93, -0·78] |
| 3_D | Single_ST | -0·47 | [-1·36, 0·43] |
| 4_A | Doesnt_have_CBT | -1·20 | [-1·61, -0·78] |
| 4_A | Has_CBT | -1·60 | [-2·13, -1·08] |
| 4_B | Doesnt_have_HA | -1·14 | [-1·81, -0·46] |
| 4_B | Has_HA | -1·59 | [-2·19, -0·98] |
| 4_C | Doesnt_have_SC | -1·28 | [-1·69, -0·87] |
| 4_C | Has_SC | -1·46 | [-1·98, -0·94] |
| 4_D | Doesnt_have_ST | -1·56 | [-1·99, -1·13] |
| 4_D | Has_ST | -1·09 | [-1·58, -0·61] |
| 5 | Brain | -1·47 | [-2·02, -0·92] |
| 5 | Brain_and_Ear | -1·50 | [-2·05, -0·94] |
| 5 | Ear | -1·07 | [-1·62, -0·52] |

*Note.* Values depict least-squares mean changes with 95% CI in square brackets. Objective 1: Single vs. Combination (all treatments); Objective 2: All treatments against each other; Objective 3: Single vs. Combination (separately); Objective 4: Treatments with vs. treatments without; Objective 5: Ear-mediated vs. brain-mediated vs. ear- and brain-mediated treatments. CBT = Cognitive-Behavioural Therapy; HA = Hearing Aids; SC = Structured Counselling; ST = Sound Therapy.

40. ***Table S30: Secondary outcome (NRS 5: “How easy is it for you to ignore your tinnitus at present?”) at final visit***

| **Objective** | **Contrast** | **Change from baseline** | **95% CI** |
| --- | --- | --- | --- |
| 1 | Combination | -1·71 | [-2·26, -1·16] |
| 1 | Single | -1·20 | [-1·74, -0·66] |
| 2 | CBT | -1·67 | [-2·82, -0·52] |
| 2 | CBT + HA | -2·41 | [-4·51, -0·30] |
| 2 | CBT + SC | -1·92 | [-3·12, -0·73] |
| 2 | CBT + ST | -1·75 | [-2·88, -0·63] |
| 2 | HA | -1·29 | [-2·33, -0·25] |
| 2 | HA + SC | -0·87 | [-2·73, 0·98] |
| 2 | HA + ST | -2·18 | [-3·69, -0·66] |
| 2 | SC | -1·18 | [-2·27, -0·10] |
| 2 | SC + ST | -1·36 | [-2·39, -0·32] |
| 2 | ST | -0·68 | [-1·72, 0·36] |
| 3_A | Combi_CBT | -1·92 | [-2·71, -1·12] |
| 3_A | Single_CBT | -1·67 | [-2·84, -0·50] |
| 3_B | Combi_HA | -1·85 | [-2·88, -0·81] |
| 3_B | Single_HA | -1·29 | [-2·33, -0·25] |
| 3_C | Combi_SC | -1·50 | [-2·22, -0·79] |
| 3_C | Single_SC | -1·18 | [-2·26, -0·11] |
| 3_D | Combi_ST | -1·66 | [-2·35, -0·97] |
| 3_D | Single_ST | -0·68 | [-1·75, 0·39] |
| 4_A | Doesnt_have_CBT | -1·21 | [-1·69, -0·74] |
| 4_A | Has_CBT | -1·84 | [-2·49, -1·19] |
| 4_B | Doesnt_have_HA | -0·95 | [-1·73, -0·16] |
| 4_B | Has_HA | -1·58 | [-2·29, -0·87] |
| 4_C | Doesnt_have_SC | -1·49 | [-1·99, -0·99] |
| 4_C | Has_SC | -1·41 | [-2·01, -0·80] |
| 4_D | Doesnt_have_ST | -1·52 | [-2·05, -0·99] |
| 4_D | Has_ST | -1·37 | [-1·94, -0·81] |
| 5 | Brain | -1·58 | [-2·26, -0·90] |
| 5 | Brain_and_Ear | -1·55 | [-2·22, -0·88] |
| 5 | Ear | -1·21 | [-1·87, -0·55] |

*Note.* Values depict least-squares mean changes with 95% CI in square brackets. Objective 1: Single vs. Combination (all treatments); Objective 2: All treatments against each other; Objective 3: Single vs. Combination (separately); Objective 4: Treatments with vs. treatments without; Objective 5: Ear-mediated vs. brain-mediated vs. ear- and brain-mediated treatments. CBT = Cognitive-Behavioural Therapy; HA = Hearing Aids; SC = Structured Counselling; ST = Sound Therapy.

41. ***Table S31: Secondary outcome (NRS 6: “How unpleasant is your tinnitus at present?”) at final visit***

| **Objective** | **Contrast** | **Change from baseline** | **95% CI** |
| --- | --- | --- | --- |
| 1 | Combination | -1·37 | [-1·83, -0·91] |
| 1 | Single | -1·16 | [-1·61, -0·71] |
| 2 | CBT | -1·31 | [-2·31, -0·31] |
| 2 | CBT + HA | -1·93 | [-3·64, -0·23] |
| 2 | CBT + SC | -1·93 | [-2·90, -0·97] |
| 2 | CBT + ST | -1·11 | [-2·06, -0·16] |
| 2 | HA | -1·40 | [-2·24, -0·55] |
| 2 | HA + SC | -1·32 | [-2·81, 0·16] |
| 2 | HA + ST | -1·38 | [-2·61, -0·15] |
| 2 | SC | -1·14 | [-2·05, -0·24] |
| 2 | SC + ST | -1·00 | [-1·83, -0·16] |
| 2 | ST | -0·80 | [-1·65, 0·05] |
| 3_A | Combi_CBT | -1·57 | [-2·22, -0·92] |
| 3_A | Single_CBT | -1·31 | [-2·31, -0·31] |
| 3_B | Combi_HA | -1·51 | [-2·39, -0·64] |
| 3_B | Single_HA | -1·40 | [-2·29, -0·50] |
| 3_C | Combi_SC | -1·40 | [-1·98, -0·82] |
| 3_C | Single_SC | -1·14 | [-2·04, -0·25] |
| 3_D | Combi_ST | -1·11 | [-1·70, -0·52] |
| 3_D | Single_ST | -0·80 | [-1·67, 0·07] |
| 4_A | Doesnt_have_CBT | -1·13 | [-1·52, -0·73] |
| 4_A | Has_CBT | -1·49 | [-2·03, -0·95] |
| 4_B | Doesnt_have_HA | -1·01 | [-1·67, -0·36] |
| 4_B | Has_HA | -1·46 | [-2·05, -0·86] |
| 4_C | Doesnt_have_SC | -1·22 | [-1·64, -0·81] |
| 4_C | Has_SC | -1·33 | [-1·82, -0·83] |
| 4_D | Doesnt_have_ST | -1·46 | [-1·89, -1·03] |
| 4_D | Has_ST | -1·02 | [-1·49, -0·55] |
| 5 | Brain | -1·45 | [-2·01, -0·89] |
| 5 | Brain_and_Ear | -1·18 | [-1·74, -0·62] |
| 5 | Ear | -1·15 | [-1·69, -0·61] |

*Note*. Values depict least-squares mean changes with 95% CI in square brackets. Objective 1: Single vs. Combination (all treatments); Objective 2: All treatments against each other; Objective 3: Single vs. Combination (separately); Objective 4: Treatments with vs. treatments without; Objective 5: Ear-mediated vs. brain-mediated vs. ear- and brain-mediated treatments. CBT = Cognitive-Behavioural Therapy; HA = Hearing Aids; SC = Structured Counselling; ST = Sound Therapy.

42. ***Figure S11: Per protocol (N = 185) Primary outcome (THI)***

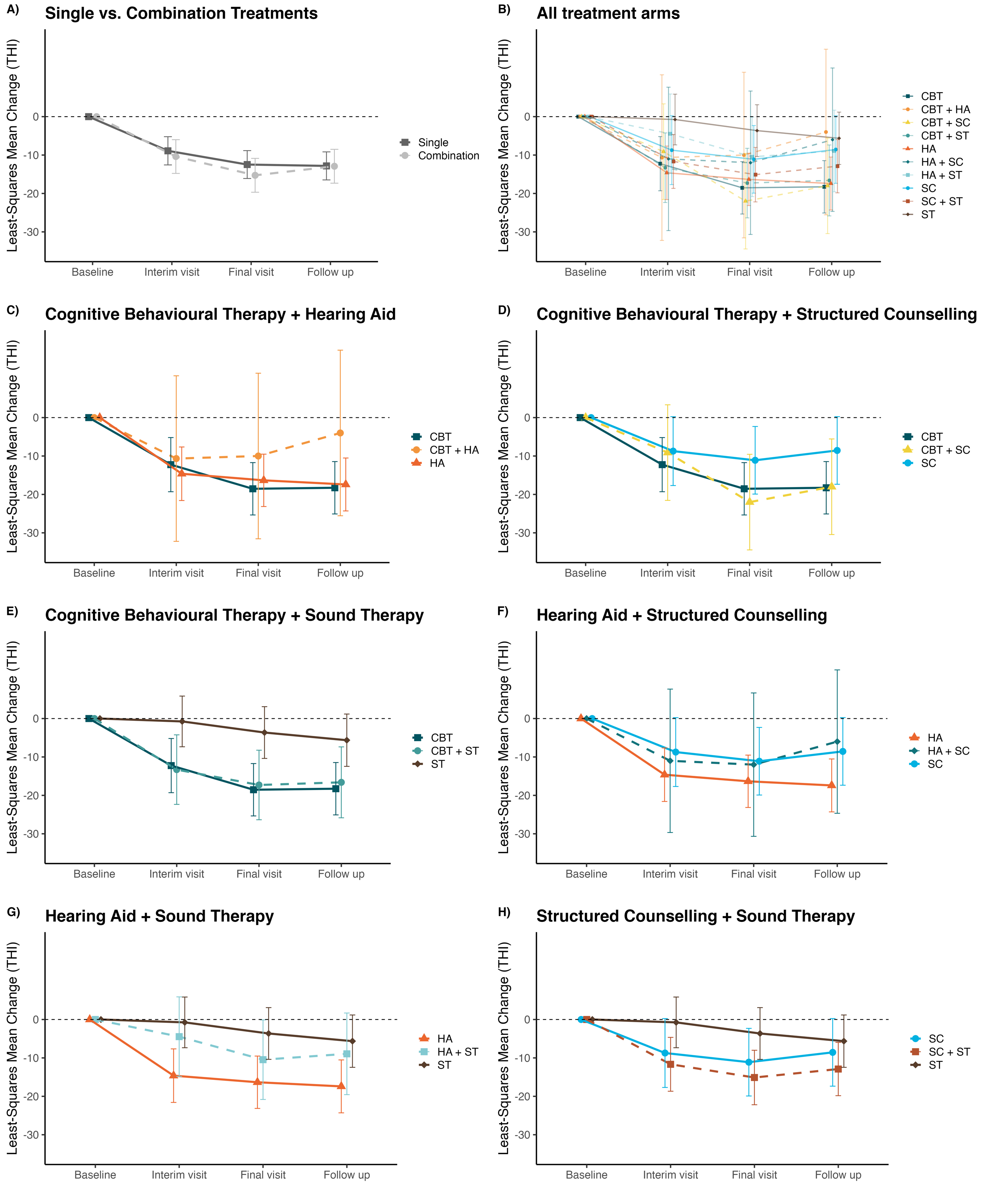

*Note.* Change from Baseline in THI total score to interim visit (6w), final visit (12w) and follow-up (36w) for A) single and combination treatments; B) all treatments; C) CBT + HA; D) CBT + SC; E) CBT + ST; F) HA + SC; G) HA + ST; and H) SC + ST. Total THI scores range from 0 to 100, with higher scores indicating greater severity of tinnitus. Error bars represent 95% confidence intervals.

43. ***Table S32: Per protocol (N = 185) Primary outcome (THI) at Final Visit***

| **Objective** | **Contrast** | **Change from baseline** | **95% CI** |
| --- | --- | --- | --- |
| 1 | Combination | -15·27 | [-19·68, -10·86] |
| 1 | Single | -12·47 | [-16·10, -8·84] |
| 2 | CBT | -18·53 | [-25·35, -11·72] |
| 2 | CBT + HA | -10·00 | [-31·55, 11·55] |
| 2 | CBT + SC | -22·00 | [-34·44, -9·56] |
| 2 | CBT + ST | -17·29 | [-26·35, -8·24] |
| 2 | HA | -16·33 | [-23·15, -9·52] |
| 2 | HA + SC | -12·00 | [-30·67, 6·67] |
| 2 | HA + ST | -10·46 | [-20·82, -0·11] |
| 2 | SC | -11·11 | [-19·91, -2·31] |
| 2 | SC + ST | -15·10 | [-22·19, -8·01] |
| 2 | ST | -3·64 | [-10·38, 3·10] |
| 3_A | Combi_CBT | -18·00 | [-25·29, -10·71] |
| 3_A | Single_CBT | -18·53 | [-25·70, -11·36] |
| 3_B | Combi_HA | -10·70 | [-18·27, -3·13] |
| 3_B | Single_HA | -16·33 | [-22·52, -10·15] |
| 3_C | Combi_SC | -16·36 | [-22·06, -10·66] |
| 3_C | Single_SC | -11·11 | [-19·68, -2·54] |
| 3_D | Combi_ST | -14·70 | [-19·71, -9·69] |
| 3_D | Single_ST | -3·63 | [-10·51, 3·24] |
| 4_A | Doesnt_have_CBT | -11·38 | [-14·75, -8·00] |
| 4_A | Has_CBT | -18·27 | [-23·16, -13·38] |
| 4_B | Doesnt_have_HA | -10·84 | [-16·53, -5·15] |
| 4_B | Has_HA | -14·08 | [-19·28, -8·88] |
| 4_C | Doesnt_have_SC | -13·06 | [-16·46, -9·66] |
| 4_C | Has_SC | -14·76 | [-19·69, -9·82] |
| 4_D | Doesnt_have_ST | -16·19 | [-20·08, -12·31] |
| 4_D | Has_ST | -10·86 | [-14·86, -6·86] |
| 5 | Brain | -16·74 | [-21·73, -11·75] |
| 5 | Brain_and_Ear | -15·30 | [-20·54, -10·06] |
| 5 | Ear | -9·98 | [-14·37, -5·59] |

*Note.* Values depict least-squares mean changes with 95% CI in square brackets. Objective 1: Single vs. Combination (all treatments); Objective 2: All treatments against each other; Objective 3: Single vs. Combination (separately); Objective 4: Treatments with vs. treatments without; Objective 5: Ear-mediated vs. brain-mediated vs. ear- and brain-mediated treatments. CBT = Cognitive-Behavioural Therapy; HA = Hearing Aids; SC = Structured Counselling; ST = Sound Therapy.

44. ***Table S33: Per Protocol 2 (N = 155) Primary Outcome (THI) at Final Visit***

| **Objective** | **Contrast** | **Change from baseline** | **95% CI** |
| --- | --- | --- | --- |
| 1 | Combination | -15·49 | [-20·31, -10·68] |
| 1 | Single | -12·47 | [-16·54, -8·40] |
| 2 | CBT | -19·05 | [-27·26, -10·83] |
| 2 | CBT + HA | -10·00 | [-31·74, 11·74] |
| 2 | CBT + SC | -22·75 | [-36·06, -9·44] |
| 2 | CBT + ST | -18·29 | [-28·35, -8·22] |
| 2 | HA | -17·48 | [-24·73, -10·23] |
| 2 | HA + SC | -15·00 | [-41·63, 11·63] |
| 2 | HA + ST | -9·64 | [-20·99, 1·72] |
| 2 | SC | -10·38 | [-19·79, -0·96] |
| 2 | SC + ST | -14·84 | [-22·26, -7·41] |
| 2 | ST | -3·07 | [-10·55, 4·40] |
| 3_A | Combi_CBT | -18·72 | [-26·52, -10·92] |
| 3_A | Single_CBT | -19·05 | [-27·56, -10·53] |
| 3_B | Combi_HA | -10·38 | [-19·03, -1·72] |
| 3_B | Single_HA | -17·48 | [-24·15, -10·82] |
| 3_C | Combi_SC | -16·62 | [-22·91, -10·34] |
| 3_C | Single_SC | -10·38 | [-19·76, -0·99] |
| 3_D | Combi_ST | -14·66 | [-20·15, -9·16] |
| 3_D | Single_ST | -3·05 | [-10·82, 4·72] |
| 4_A | Doesnt_have_CBT | -11·52 | [-15·20, -7·84] |
| 4_A | Has_CBT | -18·87 | [-24·49, -13·25] |
| 4_B | Doesnt_have_HA | -11·03 | [-17·36, -4·71] |
| 4_B | Has_HA | -14·84 | [-20·60, -9·07] |
| 4_C | Doesnt_have_SC | -13·22 | [-17·04, -9·41] |
| 4_C | Has_SC | -14·73 | [-20·07, -9·38] |
| 4_D | Doesnt_have_ST | -16·62 | [-20·97, -12·28] |
| 4_D | Has_ST | -10·81 | [-15·19, -6·44] |
| 5 | Brain | -16·62 | [-22·33, -10·91] |
| 5 | Brain_and_Ear | -15·63 | [-21·36, -9·90] |
| 5 | Ear | -10·32 | [-15·14, -5·51] |

*Note.* Values depict least-squares mean changes with 95% CI in square brackets. Per protocol 2: Additional exclusion of patients where time frame between baseline and final visit exceeded 18 weeks. Objective 1: Single vs. Combination (all treatments); Objective 2: All treatments against each other; Objective 3: Single vs. Combination (separately); Objective 4: Treatments with vs. treatments without; Objective 5: Ear-mediated vs. brain-mediated vs. ear- and brain-mediated treatments. CBT = Cognitive-Behavioural Therapy; HA = Hearing Aids; SC = Structured Counselling; ST = Sound Therapy.

45. ***Table S34: Sensitivity Analysis - Primary outcome (THI) at Final Visit***

| **Objective** | **Contrast** | **Change from baseline** | **95% CI** |
| --- | --- | --- | --- |
| 1 | Combination | -15·10 | [-17·96, -12·24] |
| 1 | Single | -11·53 | [-14·34, -8·72] |
| 2 | CBT | -18·12 | [-24·10, -12·14] |
| 2 | CBT + HA | -16·00 | [-27·17, -4·82] |
| 2 | CBT + SC | -19·17 | [-25·55, -12·79] |
| 2 | CBT + ST | -14·02 | [-19·96, -8·08] |
| 2 | HA | -14·50 | [-19·74, -9·27] |
| 2 | HA + SC | -20·67 | [-29·92, -11·42] |
| 2 | HA + ST | -12·92 | [-20·59, -5·25] |
| 2 | SC | -11·66 | [-17·35, -5·97] |
| 2 | SC + ST | -12·48 | [-17·77, -7·19] |
| 2 | ST | -3·03 | [-8·39, 2·33] |
| 3_A | Combi_CBT | -16·36 | [-20·49, -12·23] |
| 3_A | Single_CBT | -18·31 | [-24·41, -12·21] |
| 3_B | Combi_HA | -15·98 | [-20·92, -11·04] |
| 3_B | Single_HA | -14·54 | [-19·50, -9·58] |
| 3_C | Combi_SC | -16·02 | [-19·74, -12·30] |
| 3_C | Single_SC | -11·66 | [-17·35, -5·96] |
| 3_D | Combi_ST | -13·05 | [-16·66, -9·44] |
| 3_D | Single_ST | -3·01 | [-8·52, 2·51] |
| 4_A | Doesnt_have_CBT | -11·40 | [-13·90, -9·00] |
| 4_A | Has_CBT | -16·90 | [-20·30, -13·50] |
| 4_B | Doesnt_have_HA | -11·09 | [-15·19, -7·00] |
| 4_B | Has_HA | -15·27 | [-18·92, -11·61] |
| 4_C | Doesnt_have_SC | -12·31 | [-14·91, -9·71] |
| 4_C | Has_SC | -14·70 | [-17·87, -11·54] |
| 4_D | Doesnt_have_ST | -15·97 | [-18·68, -13·27] |
| 4_D | Has_ST | -10·06 | [-13·02, -7·09] |
| 5 | Brain | -16·00 | [-19·49, -12·50] |
| 5 | Brain_and_Ear | -14·43 | [-17·92, -10·94] |
| 5 | Ear | -9·69 | [-13·09, -6·29] |

*Note.* Values depict least-squares mean changes with 95% CI in square brackets. Sensitivity analysis for the primary outcome was performed without imputation of the primary outcome.
Objective 1: Single vs. Combination (all treatments); Objective 2: All treatments against each other; Objective 3: Single vs. Combination (separately); Objective 4: Treatments with vs. treatments without; Objective 5: Ear-mediated vs. brain-mediated vs. ear- and brain-mediated treatments. CBT = Cognitive-Behavioural Therapy; HA = Hearing Aids; SC = Structured Counselling; ST = Sound Therapy.

46. **References**

Probst T, Pryss RC, Langguth B, Spiliopoulou M, Landgrebe M, Vesala M, Harrison S, Schobel J, Reichert M, Stach M and Schlee W. Outpatient tinnitus clinic, self-help web platform, or mobile application to recruit tinnitus study samples? *Frontiers in Aging Neuroscience* 2017; *9*: 113.

47. **Published Study Protocol & Statistical Analysis Plan**

**Study Protocol**

Schoisswohl S, Langguth B, Schecklmann M, *et al.* Unification of Treatments and Interventions for Tinnitus Patients (UNITI): a study protocol for a multi-center randomized clinical trial. *Trials* 2021; 22: 875.

<https://trialsjournal.biomedcentral.com/articles/10.1186/s13063-021-05835-z>

**Statistical Analysis Plan**

Simoes JP, Schoisswohl S, Schlee W, *et al.* The statistical analysis plan for the unification of treatments and interventions for tinnitus patients randomized clinical trial (UNITI-RCT). *Trials* 2023; 24: 472

<https://trialsjournal.biomedcentral.com/articles/10.1186/s13063-023-07303-2>
